# Non-pharmacological interventions on depression among eye disease patients: A systematic review

**DOI:** 10.1101/2025.10.07.25337378

**Authors:** Jason Sheng, Michael Huang, Amardeep Thind, Monali S. Malvankar-Mehta

**Author notes:** These authors contributed equally to this work. These authors contributed equally to this work (Michael Huang, Amardeep Thind, Monali S. Malvankar-Mehta).

## Abstract

1

**Background:** The objective of the current systematic review is to synthesize and qualitatively summarize data with regards to the effectiveness of non-pharmacological therapies in treating depression among visually impaired older adults.

**Methods:** MEDLINE, EMBASE, Cochrane Library, and CINAHL were initially searched on March 24, 2023, whereas Web of Science and all conference databases were searched on May 30, 2023. All databases were searched once again on June 16, 2025. Studies were uploaded to Covidence and following duplicate removal, 3509 studies proceeded to title and abstract screening. Studies that investigated the impact of non-pharmacological interventions to adults (aged 18 years old) with eye disease on depression were included. Studies administering both pharmacological therapies alone and in combination with non-pharmacological therapies were excluded. Review articles, editorials, case reports, and case series were excluded. Study quality was assessed using The Tool to Assess Risk of Bias in Randomized Controlled Trials, Tool to Assess Risk of Bias in Cohort Studies, and the Risk of Bias in Non-Randomized Studies – of Interventions (ROBINS-I). Following risk of bias assessment, study data were extracted and narratively synthesized.

**Results:** A total of 30 full-text articles were included. Outcome data such as depression score at each measurement timepoint, change in depression score at each measurement timepoint, effect measures for depression, and regression data on depression score were extracted and qualitatively analyzed.

**Conclusion:** Problem solving treatment (PST) appears to offer short term relief for depression, but further studies should be conducted to investigate the necessity of booster treatments to maintain long-term effectiveness. Physical activity shows promise in improving depression outcomes among visually impaired older adults, however, additional studies with stronger evidence from randomized control trials controlling for the amount and type of exercise are necessary before reaching a firm conclusion.

## 1.1 Introduction

According to the Canadian Network for Mood and Anxiety Treatments (CANMAT), non-pharmacological interventions such as psychotherapy, exercise, or complementary and alternative medicines can be administered alone or in conjunction with antidepressants among milder cases of depression (1). Adjunctive treatment including antidepressants and psychotherapy are recommended for severe cases of depression (1). Electroconvulsive therapy should be administered to life-threatening cases of depression (1). Eye disease patients in particular, experience a unique set of circumstances that complicates the treatment of depression. More specifically, eye disease patients likely suffer from comorbidities requiring the treatment with multiple medications (2,3) which makes them vulnerable to drug-to-drug and drug-to-illness interactions (4). Many of these patients may consequently be precluded from pharmacological treatments for depression. There is also a substantial amount of research demonstrating a relationship between antidepressant use and the exacerbation of eye disease (5–15). As a result, further research in non-pharmacological interventions for the treatment of depression among eye disease is warranted.

There are several nuances when prescribing standard of care pharmacological treatment for depression among older adults with a physical health problem. Older adults with a physical health problem are likely to be taking a number of medications to address comorbidities (2,3). As a result, they are at a greater risk of suffering from drug to drug as well as drug to illness interactions while undergoing pharmacological treatment for depression (4). Patients with physical health problems have also commonly been excluded from meta-analyses examining the effectiveness of pharmacological treatments (16). In addition, up to 63% of patients that use antidepressants experience adverse events (17). Ultimately, this results in treatment discontinuation among 7% to 15% of patients that are prescribed antidepressants (17). Taken together, antidepressants are discouraged among this subpopulation.

Although there has been research demonstrating that non-pharmacological interventions may be effective in the treatment of depression among older adults (18–21), there is yet to be a study to examine the effectiveness of these treatments among visually impaired older adults in particular. Considering this, the objective of the current systematic review is to synthesize data examining the effectiveness of non-pharmacological therapies in treating depression among visually impaired older adults.

## 1.2 Materials and methods

### 1.2.1 Data sources and searches

The current systematic review was conducted based on the Preferred Reporting Items for Systematic Reviews and Meta-Analyses (PRISMA) statements (22) and as outlined in a registered protocol (PROSPERO 2023: CRD42023451502). Databases including MEDLINE, EMBASE, Cochrane Library, CINAHL, and Web of Science were searched for relevant articles. The following conferences were searched as well: Canadian Ophthalmological Society, American Academy of Ophthalmology, and Association for Vision in Research and Ophthalmology. Backward citation searching was conducted while performing background research to find potentially relevant articles. There were no restrictions placed on the language or publication date of articles.

The search strategy combined search terms for depression with search terms for some of the most prevalent eye diseases (23) and search terms for non-pharmacological interventions that have been commonly studied in previous reviews (24,25). The complete search strategy can be found in Appendix 1.

### 1.2.2 Inclusion/exclusion criteria

Studies enrolling adults (aged at least 18 years old) with some of the most prevalent eye diseases (23) including diabetic retinopathy, AMD, cataracts, glaucoma, optic nerve disease, and uveitis were included. The interventions of interest included those that have been studied in similar reviews in the past (24,25) such as meditation, behavioral therapy, psychotherapy, physical activity, tai-chi, acupuncture, herbal medicine, and nutritional supplements. Studies administering both pharmacological therapies alone and in combination with non-pharmacological therapy were excluded. There was no restriction placed on the comparator administered. Outcomes such as depression score at each measurement timepoint, change in depression score at each measurement timepoint, regression data modelling depression outcomes, and effect measures for depression were included. Experimental and cohort studies were included, whereas review articles, editorials, case reports, and case series were excluded.

Studies that fulfilled the inclusion/exclusion criteria were imported into Covidence systematic review software (26). Duplicates were automatically removed upon study importation to Covidence. Following this, systematic screening was conducted by two independent reviewers (J.S. and M.H.). Level 1 screening included title and abstract screening. The reviewers were instructed to include articles that administered non-pharmacological interventions to eye disease patients for the treatment of depression. Furthermore, review articles, editorials, case reports, and case series were excluded. All other study designs were included. Level 2 screening included full-text screening where reviewers were instructed to include articles that enrolled adult patients. Cohen’s kappa coefficient and the raw percentage of agreement between the two reviewers were calculated at each stage before conflict resolution. Conflicts were resolved by discussion between the reviewers until a consensus was reached. If a consensus could not be reached, the conflict was brought to the attention of a third reviewer (M.M.).

### 1.2.3 Data extraction and analysis

Data extraction was conducted by a single reviewer (J.S.) and checked by two other reviewers (A.T. and M.M.) for accuracy. Study data such as first author, year of publication, geographical location of study, study design, intervention and comparator administered, sample size, questionnaire administered, mode of questionnaire administration, and measurement timepoint(s) were extracted. Participant characteristics such as sex, age, eye disease, history of depression, and visual acuity were also extracted. Outcome data were extracted as well.

The Risk of Bias assessment tools by CLARITY Group of McMaster University (27), Instrument for Cross-Sectional Surveys of Attitudes and Practices, Tool to Assess Risk of Bias in Cohort Studies, Tool to Assess Risk of Bias in Randomized Controlled Trials, and Tool to Assess Risk of Bias in Longitudinal Symptom Research were used to assess risk of bias based on the study design of the included study. Risk of bias assessment was conducted by a single reviewer (J.S.).

A meta-analysis is appropriate if there is overlap in the patients, interventions, and outcomes investigated across included studies (28). Given the heterogeneity of the current dataset, a meta-analysis would result in unreliable pooled estimates. In addition, there was a wide range of effect measures reported across studies which could not be pooled together. As a result, the application of a meta-analysis as described in the study protocol was not possible. Alternatively, data may be organized into homogenous groups to enable narrative synthesis of results (29). Existing reviews have narratively synthesized data according to overlapping characteristics in the patient, intervention, comparator, and outcome (PICO) framework (30–32). Given the aforementioned challenges, the results for the effectiveness of non-pharmacological interventions were narratively synthesized according to similarities across dimensions of the PICO framework.

## 1.3 Results

### 1.3.1 Search strategy

Initially, MEDLINE, EMBASE, Cochrane Library, and CINAHL were searched on March 24, 2023, whereas Web of Science and all conference databases were searched on May 30, 2023. All databases were searched once again on June 16, 2025. The search strategy yielded a total of 6678 studies that were uploaded to Covidence (26). The gray literature search yielded 1 study from the American Academy of Ophthalmology. After performing backward citation searching while conducting background research, 3 potentially relevant studies were retrieved from Holloway et al. (2015) and another 7 were retrieved from van der Aa et al. (2016) (33). These studies were also uploaded to Covidence. Following duplicate removal, 3509 articles remained. A total of 3439 studies were deemed irrelevant after title and abstract screening because the investigators did not examine non-pharmacological treatment among eye disease patients for the treatment of depression. Following this, 70 studies proceeded to full-text review. After full-text review, 40 studies were excluded, 23 of which employed the wrong study design, 7 studied the wrong outcome, 3 studied the wrong patient population, 1 was an animal study, and 6 were irretrievable. Finally, 30 studies were included for qualitative analysis. The PRISMA flowchart as well as the reasons for exclusion are depicted in Fig 1. The Cohen’s kappa statistic for title and abstract screening was 0.6 indicating moderate agreement. The Cohen’s kappa statistic for full-text review was 0.9, indicating excellent agreement.

**Fig 1.** PRISMA flowchart of the study selection process

### 1.3.2 Characteristics of included studies

Study and participant characteristics are reported in Tables 1 and 2. Although the search strategy limited studies to participants that were at least 18 years old, the participants in the included studies were commonly older adults due to the increasing risk of eye disease associated with increasing age. In total, 11 of the 30 included studies were conducted in the USA,(34–44) 5 in Australia (45–47), 2 in Germany (48,49), 4 in China (50–53), and 1 each in Norway (54), Canada (55), England (56), Japan (57), the Netherlands and Belgium (58), the United Kingdom (59), Egypt (60), and Hong Kong (61). There were 21 randomized control trials (RCT) (34,35,38–40,42–45,47,49–51,55,56,58,59,62–64), 4 single-arm trials (36,37,46,48), 2 quasi-experimental studies (57), and 2 prospective cohort studies (41,54), and 1 retrospective cohort study (53). With regards to the intervention or exposure of interest, 5 examined PST (43,44,46,56,63,64), 4 examined SM (38,39,47,62), 4 examined physical activity (36,37,54,59), and 1 study each examined low vision rehabilitation (LVR) (48), behavioral activation (BA) and LVR (35), psychoeducational group therapy and LVR (34), optical and adaptive device use (41), rational emotive behavior therapy (REBT) (42), skills training with group counselling and individual therapy (57), emotion or problem-focused therapy (49), CBT (53), psychological and physical therapy (50), mindfulness based music therapy (61), alternate nostril breathing exercises (60), written and audio tools incorporating cognitive-behavioral principles in addition to phone calls from a coach (55), motivational psychological nursing as well as cognitive behavioral interventions and routine nursing (51), expressive writing (40), the Alexander technique (45), stepped care (58), and acupuncture (52). Brief descriptions of the interventions/exposures investigated in the included studies can be found in Appendix 2.

**Table 1.** Characteristics of Included Studies.

| **First Author and Year of Publication** | **Country** | **Study Design** | **Intervention or exposure arm** | **Comparator arm** | **Sample Size** |
| --- | --- | --- | --- | --- | --- |
| Adams 2019 | United Kingdom | RCT | Exercise program | Usual activities (no intervention) | trt=1: 33<br>trt=0: 31 |
| Bernbaum 1988/1989<sup>a</sup> | USA | Single arm trial | Exercise protocol and patient education sessions | Not applicable | 29 |
| Brody 1999 | USA | RCT | SM | Wait-list | trt=1: 44<br>trt=0: 48 |
| Brody 2006 | USA | RCT | SM | Tape recorded health education program or wait-list | trt=1: 12<br>trt=0: 20 |
| Brunes 2017 | Norway | Prospective cohort | Physical activity | Not applicable | 34,393 |
| Bryan 2016 | USA | RCT | Expressive writing | Neutral writing | trt=1: 38<br>trt=0: 43 |
| Chan 2023 | Hong Kong | RCT | Mindfulness based music therapy or mindfulness intervention | Waitlist control group | Mindfulness based music therapy: 31<br>Mindfulness intervention: 30<br>Waitlist control group: 31 |
| Duan 2024 | China | Quasi-experimental | Acupuncture | Sodium hyaluronate | trt=1: 27<br>trt=0: 21 |
| Girdler 2010 | Australia | RCT | SM | Usual care (one-on-one case management model, initial interview with patient to discuss service needs, vision assessment and prescription of vision aids, and referral to service provider) | trt=1: 36<br>trt=0: 41 |
| Gleeson 2017 | Australia | RCT | The Alexander technique and usual care | Usual care (access to orientation and mobility programs) | trt=1: 60<br>trt=0: 60 |
| Holloway 2018 | Australia | Single arm trial | PST | Not applicable | 55 |
| Horowitz 2006 | USA | Prospective cohort | Optical and adaptive device use | Not applicable | 438 |
| Ismail 2023 | Egypt | RCT | Alternate nostril breathing exercise | Waitlist control group | trt=1: 30<br>trt=0: 30 |
| Jalali 2014 | USA | RCT | REBT | NR | trt=1: 30<br>trt=0: 30 |
| Kamga 2017 | Canada | RCT | Large print written and audio tools incorporating cognitive-behavioral principles plus three 10-minute telephone calls from a lay coach | Usual care | trt=1: 41<br>trt=0: 39 |
| Lin 2025 | China | Retrospective cohort | CBT | Participants not treated by CBT | trt=1: 859<br>trt=0: 2151 |
| Nollett 2016 | England | RCT | PST | Referral to patient's physician or waiting list control | trt=1: 24<br>trt=0: 61 |
| Rees 2015 | Australia | RCT | SM and usual care | Usual care (Initial assessment by a multidisciplinary team from a LVR service, visual assessment, prescription of vision aids, and training, as requested) | trt=1: 93<br>trt=0: 60 |
| Rees 2017 | Australia | RCT | PST and brochures | Brochures discussing diabetes-specific topics and usual care | trt=1: 21<br>trt=0: 19 |
| Renieri 2013 | Germany | Single arm trial | LVR | Not applicable | 87 |
| Rovner 2007 | USA | RCT | PST and usual care | Usual care | trt=1: 105<br>trt=0: 101 |
| Rovner 2013 | USA | RCT | PST | ST | trt=1: 121<br>trt=0: 120 |
| Rovner 2014 | USA | RCT | BA and LVR | ST and LVR | trt=1: 96<br>trt=0: 92 |
| Sun 2012 | China | RCT | Psychological and physical therapy | Physical therapy | trt=1: 50<br>trt=0: 50 |
| Trozzolino 2003 | USA | RCT | Group counseling including CBT and patient education in addition to LVR | LVR | trt=1: 24<br>trt=0: 24 |
| Ueda 2013 | Japan | Quasi-experimental | Skills training with group counseling and individual therapy | Skills training | trt=1: 37<br>trt=0: 42 |
| van der Aa 2015 | Netherlands and Belgium | RCT | Stepped care and usual care | Usual care (LVR and/or care that was provided by other healthcare providers) | trt=1: 131<br>trt=0: 134 |
| Wahl 2006 | Germany | RCT | Emotion-focused or problem-focused therapy | NR | trt=1: 45<br>trt=0: 22 |
| Zhu 2019 | China | RCT | Motivational psychological nursing and cognitive behavioral interventions and routine nursing | Routine nursing | trt=1: 39<br>trt=0: 39 |

Notes: <sup>a</sup>: The study population is the same, but Bernbaum et al. reported the results in 2 separate papers; trt=1: Intervention or exposure arm; trt=0: Comparator arm; ST: Supportive therapy; NR: Not reported

**Table 2.** Characteristics of the Participants of the Included Studies.

| First Author and Year of Publication | Male: n (%) | Age (Years) | Eye Disease: n (%) | Baseline depression | Visual Acuity |
| --- | --- | --- | --- | --- | --- |
| Adams 2019 | trt=1: 14 (42)<br>trt=0: 11 (35) | trt=1: mean±SD (range):<br>79.3±8.7 (61 to 95)<br>trt=0: mean±SD (range):<br>76.5±9.7 (62 to 95) | NR | NR | Visually impaired<br>trt=1: 33 (100)<br>trt=0: 31 (100) |
| Bernbaum 1988 | 11 (37.9) | median (range): 38.0 (21.0 to 66.0) | Diabetic retinopathy: 29 (100.0) | NR | <20/100: 29 (100.0) |
| Bernbaum 1989 | 11 (37.9) | median (range): 38.0 (21.0 to 66.0) | Visual impairment:<br><20/100: 29 (100.0) | NR | NR |
| Brody 1999 | trt=1: 17 (59.0)<br>trt=0: 10 (40.0) | trt=1: mean (SD): 78.5 (5.1)<br>trt=0: mean (SD): 79.6 (6.6) | AMD:<br>Total sample: 92 (100.0) | NR | Visual acuity (LogMAR in better eye:<br>0=20/20, 1=20/200): mean (SD):<br>trt=1: 1.0 (0.3)<br>trt=0: 1.2 (0.4)<br>p-value=0.03 |
| Brody 2006 | trt=1: 5 (41.7)<br>Tape recording group: 2 (25.0)<br>Wait-list group: 4 (33.3) | SM: mean (SD): 81.2 (9.6)<br>Tape recording: mean (SD): 81.9 (5.4)<br>Wait-list group: mean (SD): 81.6 (7.1) | AMD:<br>SM: 12 (100.0)<br>Tape recording: 8 (100.0)<br>Wait-list group: 12 (100.0) | NR | Log of best eye: mean (SD):<br>SM: 1.3 (0.5)<br>Tape recording: 1.2 (0.4)<br>Wait-list group: 1.2 (0.4)<br>p-value=0.6 |
| Brunes 2017 | 15,494 (45.0) | SRVI: mean: 51.7<br>SRNI: mean: 44.2 | NR | NR | SRVI: n (%):<br>A little: 2263 (6.6)<br>Some: 1134 (3.3)<br>Severe: 332 (0.9) |
| Bryan 2016 | Participants that completed the writing tasks: 15 (31.2)<br>Participants that completed the 3-week follow-up: 10 (27.0) | Participants that completed the writing tasks: mean±SD (range): 41.7±12.6 (20.0 to 71.0) | Stargardt's disease: 81 (100.0) | NR | NR |
| Chan 2023 | Mindfulness based music therapy: 0 (0)<br>Mindfulness intervention: 0 (0)<br>Waitlist control group: 0 (0) | Mindfulness based music therapy: 85.5 (8.6)<br>Mindfulness intervention: 88.4 (7.4)<br>Waitlist control group: 84.1 (6.5) | NR | GDS: mean (SD):<br>Mindfulness based music therapy: 10.8 (1.7)<br>Mindfulness intervention: 10.3 (1.8)<br>Waitlist control group: 10.2 (2.4)<br>DASS-21 depression domain: mean (SD):<br>Mindfulness based music therapy: 8.2 (3.1)<br>Mindfulness intervention: 7.8 (3.1)<br>Waitlist control group: 7.1 (2.8) | Blind (visual acuity of the eye $\leq 20/400$): n (%):<br>Mindfulness based music therapy: 31 (100)<br>Mindfulness intervention: 30 (100)<br>Waitlist control group: 31 (100) |
| Duan 2024 | trt=1: 4 (14.8)<br>trt=0: 5 (23.8)<br>p-value: 0.675 | trt=1: 42.0 (13.9)<br>trt=0: 41.7 (10.9)<br>p-value: 0.931 | Dry eye disease:<br>trt=1: 27 (100)<br>trt=0: 21 (100) | SDS score: mean (SD):<br>trt=1: 46.4 (11.0)<br>trt=0: 40.2 (7.7)<br>p-value: 0.086 | Corneal sensitivity:<br>trt=1: 5.7 (0.5)<br>trt=0: 5.7 (0.6)<br>p-value: 0.948 |
| Girdler 2010 | trt=1: 10 (27.8)<br>trt=0: 17 (41.5)<br>p-value=0.21 | trt=1: mean (SD): 79.4 (7.2)<br>trt=0: mean (SD): 80.4 (6.3)<br>p-value=0.53 | Visual impairment:<br>Mild:<br>trt=1: 9 (25.0)<br>trt=0: 7 (17.1)<br>Total sample: 16 (20.8)<br>Moderate:<br>trt=1: 13 (36.1)<br>trt=0: 21 (51.2)<br>Total sample: 34 (44.1)<br>Severe:<br>trt=1: 14 (38.9)<br>trt=0: 13 (31.7)<br>Total sample: 1.0 (0.5)<br>p-value=0.4 | GDS clinical threshold for depression: n (%):<br>Yes:<br>trt=1: 18 (50.0)<br>trt=0: 15 (36.6)<br>No:<br>trt=1: 18 (50.0)<br>trt=0: 26 (63.4)<br>p-value=0.24 | Log of weighted visual acuity: mean (SD):<br>trt=1: 1.0 (0.5)<br>trt=0: 1.0 (0.5)<br>p-value: 0.79 |
| Gleeson 2017 | trt=1: 17 (28.0)<br>trt=0: 18 (30.0) | trt=1: mean (SD): 74.7 (10.9)<br>trt=0: mean (SD): 74.88 (11.0) | Macular disease:<br>trt=1: 22 (37.0)<br>trt=0: 25 (42.0) | NR | Visual acuity: n (%):<br>Better than 6/18:<br>trt=1: 7 (12.0) |
|  |  |  | Glaucoma:<br>trt=1: 9 (15.0)<br>trt=0: 5 (8.0)<br>Cerebral injury:<br>trt=1: 4 (7.0)<br>trt=0: 2 (3.0)<br>Diabetic retinopathy:<br>trt=1: 3 (5.0)<br>trt=0: 3 (5.0)<br>Retinitis pigmentosa:<br>trt=1: 9 (15.0)<br>trt=0: 5 (8.0)<br>Other:<br>trt=1: 13 (22.0)<br>trt=0: 20 (33.0) |  | trt=0: 7 (12.0)<br>6/18 to 6/60:<br>trt=1: 6 (10.0)<br>trt=0: 1 (2.0)<br>6/60 or worse:<br>trt=1: 45 (75.0)<br>trt=0: 49 (82.0)<br>Peripheral loss - one eye:<br>trt=1: 4 (7.0)<br>trt=0: 4 (7.0)<br>Peripheral loss - both:<br>trt=1: 13 (22.0)<br>trt=0: 6 (10.0)<br>Central loss - one eye:<br>trt=1: 5 (8.0)<br>trt=0: 4 (7.0)<br>Total field involvement:<br>trt=1: 9 (15.0)<br>trt=0: 16 (27.0) |
| Holloway<br>2018 | 15 (27.3) | Mean (SD): 62.2 (17.6) | Macular degeneration: 14 (26.4)<br>Glaucoma: 9 (17.0)<br>Diabetic retinopathy: 10 (18.9)<br>Cataracts: 4 (7.5)<br>Other: 16 (30.2) | PHQ-9 clinical threshold for depression: n (%):<br>Yes:<br>Completed PST: 12 (66.7)<br>Withdrew: 17 (45.9)<br>Total: 29 (52.7)<br>No:<br>Completed PST: 6 (33.3)<br>Withdrew: 20 (54.1)<br>Total: 26 (47.3) | Visual acuity: n (%):<br>>6/12: 6 (11/3)<br><6/12-6/18: 16 (30.2)<br><6/18-6/60: 20 (37.7)<br><6/60: 11 (20.8) |
| Horowitz<br>2006 | 204 (46.6) | Mean (SD): 80.4 (7.4) | AMD: 305 (69.7)<br>Glaucoma: 117 (26.8)<br>Cataracts: 162 (37.1)<br>Diabetic retinopathy: 60 (13.7) | NR | Legal blindness (i.e., 20/200 or worse): 90 (20.5) |
| Ismail 2023 | trt=1: 12 (40.0)<br>trt=0: 10 (33.3) | trt=1: 69.5 (4.1)<br>trt=0: 70.0 (4.3) | Glaucoma:<br>trt=1: 30 (100)<br>trt=0: 30 (100) | HADS-D: mean (SD):<br>trt=1: 9.06 (1.46)<br>trt=0: 9.16 (1.44)<br>p=0.790 | NR |
| Jalali 2014 | NR | NR | Blindness: 60 (100.0) | NR | NR |
| Kamga 2018 | trt=1: 14 (34)<br>trt=0: 16 (41) | trt=1: mean (SD): 75.1 (11.1)<br>trt=0: mean (SD): 73 (10.8) | AMD:<br>trt=1: 22 (54.0)<br>trt=0: 22 (56.0)<br>Diabetic retinopathy:<br>trt=1: 19 (46)<br>trt=0: 17 (44) | NR | LogMAR visual acuity: median (IQR):<br>trt=1: 0.4 (0.2)<br>trt=0: 0.3 (0.2) |
| Lin 2025 | trt=1: 232 (27.0)<br>trt=0: 579 (26.9) | trt=1: mean (SD): 65.82 (5.43)<br>trt=0: mean (SD): 65.74 (5.67) | Advanced glaucoma in both eyes:<br>trt=1: 287 (33.4)<br>trt=0: 731 (34.0)<br>Advanced glaucoma in single eye:<br>trt=1: 201 (33.4)<br>trt=0: 430 (20.0)<br>Advanced cataract in both eyes:<br>trt=1: 261 (30.4)<br>trt=0: 688 (32.0)<br>Advanced cataract in single eye:<br>trt=1: 110 (12.8)<br>trt=0: 302 (14.0) | Mild to moderate depression: n (%):<br>trt=1: 859 (100.0)<br>trt=0: 2151 (100.0) | LogMAR visual acuity:<br>trt=1: 0.41 (0.2)<br>trt=0: 0.42 (0.1) |
| Nollett 2016 | PST: 8 (33.3)<br>Referral: 11 (35.5)<br>Waiting: 16 (53.3) | PST: mean (SD): 71.8 (16.7)<br>Referral: mean (SD): 67.1 (19.6)<br>Waiting: mean (SD): 72.3 (13.1) | Low vision:<br>PST: 8 (100.0)<br>Referral: 11 (100.0)<br>Waiting: 16 (100.0) | NR | NR |
| Rees 2015 | trt=1: 39 (41.9)<br>trt=0: 22 (36.7)<br>Total sample: 61 (39.9) | trt=1: mean (SD): 80.1 (8.1)<br>trt=0: mean (SD): 80.5 (8.1)<br>Total sample: 80.2 (8.1) | AMD:<br>trt=1: 65 (70.7)<br>trt=0: 41 (69.5)<br>Total sample: 106 (70.2) | NR | Presenting visual acuity (LogMAR): n (%):<br>Mild (0.34-0.48):<br>trt=1: 11 (11.8)<br>trt=0: 9 (15.0) |
|  |  |  | Glaucoma:<br>trt=1: 4 (4.4)<br>trt=0: 6 (10.2)<br>Total sample: 10 (6.6)<br>Diabetic retinopathy:<br>trt=1: 6 (6.5)<br>trt=0: 4 (6.8)<br>Total sample: 10 (6.6)<br>Other:<br>trt=1: 17 (18.5)<br>trt=0: 8 (13.6)<br>Total sample: 25 (16.6) |  | Total sample: 20 (13.1)<br>Moderate (0.5-1.0):<br>trt=1: 46 (49.5)<br>trt=0: 33 (55.0)<br>Total sample: 79 (51.6)<br>Severe (>1.04):<br>trt=1: 36 (38.7)<br>trt=0: 18 (30.0)<br>Total sample: 54 (35.3) |
| Rees 2017 | trt=1: 14 (66.7)<br>trt=0: 13 (68.4) | trt=1: mean (SD): 60.1 (7.0)<br>trt=0: mean (SD): 59.6 (8.8) | Diabetic retinopathy: 40 (100) | NR | Worst eye best presenting distance visual acuity: (LogMAR):<br>trt=1: 0.2 (0.2)<br>trt=0: 0.2 (0.4)<br>Total sample: 0.2 (0.3) |
| Renieri 2013 | 46 (52.9) | mean (SD): 74.6 (13.8) | AMD: 64 (73.6)<br>Glaucoma: 17 (19.3)<br>Diabetic retinopathy: 10 (11.5)<br>Other: 76 (88.4) | NR | Best corrected visual acuity: median (I-III quartile):<br>Study group: 20/200 (20/400-20/100)<br>Dropouts: 20/160 (20/320-20/50)<br>Overall: 20/160 (20/400-20/50)<br>p-value=0.056 |
| Rovner 2007 | trt=1: 36 (34.3)<br>trt=0: 26 (25.7) | trt=1: mean (SD): 81.3 (5.4)<br>trt=0: mean (SD): 81.0 (6.3) | AMD:<br>trt=1: 105 (100)<br>trt=0: 101 (100) | HDRS score: mean (SD):<br>trt=1: 2.10 (2.07)<br>trt=0: 2.25 (2.36) | NR |
| Rovner 2013 | trt=1: 39 (32.2)<br>trt=0: 49 (40.8) | trt=1: mean (SD): 82.7 (6.6)<br>trt=0: mean (SD): 82.8 (7.3) | AMD:<br>trt=1: 121 (100)<br>trt=0: 120 (100) | NR | Best eye, distance (LogMAR):<br>trt=1: 0.6 (0.3)<br>trt=0: 0.6 (0.3) |
| Rovner 2014 | trt=1: 26 (27.1)<br>trt=0: 30 (32.6) | trt=1: mean (SD): 85.2 (6.6)<br>trt=0: mean (SD): 82.7 (6.9) | AMD:<br>trt=1: 96 (100)<br>trt=0: 92 (100) | PHQ-9 score: mean (SD):<br>trt=1: 1.4 (2.7)<br>trt=0: 1.2 (2.3) | Best eye distance acuity (LogMAR):<br>trt=1: 0.7 (0.4)<br>trt=0: 0.7 (0.3)<br>Worst eye distance acuity (LogMAR):<br>trt=1: 1.4 (0.7)<br>trt=0: 1.4 (0.7)<br>Best eye contrast sensitivity (log CS) |
|  |  |  |  |  | trt=1: 1.5 (0.6)<br>trt=0: 1.6 (0.5)<br>Worst eye contrast sensitivity (log CS)<br>trt=1: 1.0 (0.5)<br>trt=0: 0.9 (0.5)<br>Best eye scotoma size (no. of squares)<br>trt=1: 85.7 (125.8)<br>trt=0: 109.8 (148.6)<br>Worst eye scotoma size (no. of squares)<br>trt=1: 246.6 (160.7)<br>trt=0: 249.3 (157.4) |
| Sun 2012 | NR | Mean: 62 years<br><60 years old: 15.6%<br>60-70 years old: 62.4%<br>>70 years old: 22.0% | Closed angle glaucoma: 65 (65.0)<br>Open angle glaucoma: 35 (35.0) | PHQ-9 depression score:<br>mean (SD):<br>trt=1: 5.5 (2.5)<br>trt=0: 5.6 (2.2) | NR |
| Trozzolino 2003 | trt=1: 7 (29.2)<br>trt=0: 10 (41.7) | trt=1: mean (SD): 63.5 (10.1)<br>trt=0: mean (SD): 61.4 (9.6) | Low vision (corrected visual acuity greater than 20/70 or visual fields of less than 30-40 degrees):<br>trt=1: 24 (100.0)<br>trt=0: 24 (100.0) | NR | NR |
| Ueda 2013 | trt=1: 26 (0.7)<br>trt=0: 28 (0.7) | trt=1: mean±SD (range): 43.7±11.8 (20 to 65)<br>trt=0: mean±SD (range): 47.6±13.5 (19 to 75) | Retinitis pigmentosa:<br>trt=1: 14 (37.8)<br>trt=0: 23 (54.8)<br>Optic atrophy:<br>trt=1: 8 (21.6)<br>trt=0: 2 (4.8)<br>Diabetic retinopathy:<br>trt=1: 1 (2.7)<br>trt=0: 1 (2.4)<br>Retinal detachment:<br>trt=1: 5 (13.5)<br>trt=0: 1 (2.4)<br>Glaucoma:<br>trt=1: 2 (5.4)<br>trt=0: 6 (14.3) | NR | Totally blind or light perception only:<br>trt=1: 9 (24.3)<br>trt=0: 11 (26.2)<br>Hand movement:<br>trt=1: 7 (18.9)<br>trt=0: 4 (9.5)<br>Counting finger<br>trt=1: 1 (2.7)<br>trt=0: 2 (4.8)<br>Less than 0.1<br>trt=1: 13 (35.1)<br>trt=0: 20 (47.6)<br>Greater than 0.1 but field loss:<br>trt=1: 7 (18.9)<br>trt=0: 5 (11.9) |
|  |  |  | Other or unknown causes:<br>trt=1: 7 (18.9)<br>trt=0: 9 (21.4) |  |  |
| van der Aa<br>2015 | trt=1: 40 (30.0)<br>trt=0: 40 (30.0) | trt=1: mean (SD): 72.4 (12.5)<br>trt=0: mean (SD): 74.9 (11.9) | Macular degeneration:<br>trt=1: 62 (47.0)<br>trt=0: 60 (45.0)<br>Glaucoma:<br>trt=1: 26 (20.0)<br>trt=0: 19 (14.0)<br>Cataracts:<br>trt=1: 26 (20.0)<br>trt=0: 19 (14.0)<br>Diabetic retinopathy:<br>trt=1: 5 (4.0)<br>trt=0: 10 (8.0)<br>Cerebral hemorrhage:<br>trt=1: 5 (4.0)<br>trt=0: 10 (8.0)<br>Other:<br>trt=1: 45 (34.0)<br>trt=0: 60 (45.0) | NR | LogMAR visual acuity: n (%):<br>Normal visual acuity:<br>trt=1: 9 (7.0)<br>trt=0: 15 (11.0)<br>Mild vision loss:<br>trt=1: 24 (18.0)<br>trt=0: 23 (17.0)<br>Low vision/blindness:<br>trt=1: 86 (66.0)<br>trt=0: 86 (64.0) |
| Wahl 2006 | Emotion-focused: 2 (10.0)<br>Problem-focused: 7 (32.0)<br>Comparison group: 5 (23.0) | Emotion-focused: mean±SD<br>(range): 76.5±6.8 (65.0-92.0)<br>Problem-focused: 76.6±7.1<br>(61.0-88.0)<br>Comparison group: 77.3±6.4<br>(64.0-84.0) | AMD:<br>Total sample: 67 (100.0) | NR | NR |
| Zhu 2019 | trt=1: 27 (69.2)<br>trt=0: 29 (74.4) | trt=1: mean (SD): 61.8 (5.1)<br>trt=0: mean (SD): 61.2 (5.4) | Primary glaucoma:<br>trt=1: 19 (48.7)<br>trt=0: 22 (56.4)<br>Secondary glaucoma:<br>trt=1: 12 (30.8)<br>trt=0: 10 (25.6)<br>Miscibility:<br>trt=1: 8 (20.5)<br>trt=0: 7 (17.9) | NR | NR |

Notes: GDS: Geriatric depression scale; SDS: Self rating depression scale; Hospital anxiety depression scale – Depression (HADS-D)

### 1.3.3 Problem solving treatment

According to the PICO framework, 2 of the 4 studies that administered PST overlapped in terms of the patient population, intervention, and outcome. These studies investigated the effect of PST on depression among AMD patients (43,44). Rovner et al. (2007) randomly assigned AMD patients to 6 sessions of PST delivered over the course of 8 weeks compared to usual care from their ophthalmologists and other healthcare providers (43). The odds ratio for depression as measured by the Hamilton Depression Rating Scale (HDRS) was significant at 2 months (OR: 0.4, 95% CI (0.2 to 0.9), p-value=0.03), but not at 6 months (OR: 0.7, 95% CI (0.3 to 1.4), p-value=0.29) (Table 3) (43). In addition, Rovner et al. (2007) observed that treatment group was a statistically significant predictor in a regression model plotting the odds ratio for depression diagnosis at 2 months (β=2.3, 95% CI (1.1 to 5.1)) (Table 4) (43). After activity loss as measured using the National Eye Institute Vision Function Questionnaire (NEI VFQ-17) was added to the model (β=2.6, 95% CI (1.2 to 5.5)), treatment group lost its statistical significance (β=2.0, 95% CI (0.9 to 4.5)) while activity loss remained statistically significant (43). Rovner et al. (2013) randomized AMD patients to PST or ST (44). The mean (SD) PHQ-9 score was 1.4 (2.7), 1.5 (2.8), and 1.4 (3.2) for the PST group and 1.2 (2.3), 1.3 (2.3), and 1.5 (2.9) for the ST group at baseline, 3 months, and 6 months (Table 5) (44). There was no statistically significant difference in PHQ-9 score between treatment groups at 3 months and 6 months follow-up (44).

**Table 3.**
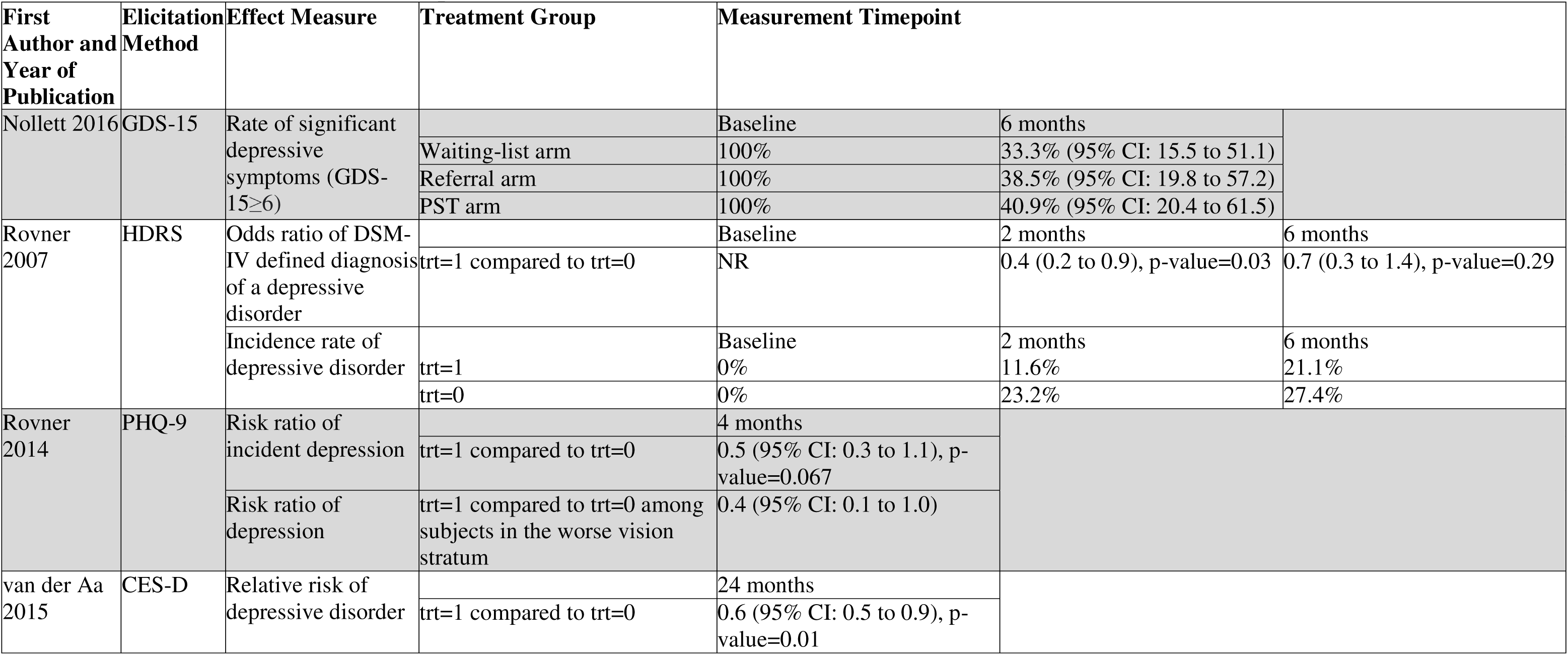
Effect Measures of Depression.

**Table 4.** Regression Equation Modelling Depression as the Outcome.

| First Author and Year of Publication | Elicitation Method | Outcome | Effect | Regression Coefficient | 95% Confidence Intervals | p-value |
| --- | --- | --- | --- | --- | --- | --- |
| Horowitz 2006 | CES-D | Short term depression | Step 1 |  |  |  |
|  |  |  | Time 1 depression | 0.6 | NR | <0.001 |
|  |  |  | Step 2 |  |  |  |
|  |  |  | Age | -0.7 | NR | >0.05 |
|  |  |  | Gender (female) | 0.005 | NR | >0.05 |
|  |  |  | Race (non-Hispanic White) | 0.06 | NR | >0.05 |
|  |  |  | Education | 0.02 | NR | >0.05 |
|  |  |  | Step 3 |  |  |  |
|  |  |  | Time 1 disability | 0.9 | NR | <0.05 |
|  |  |  | Step 4 |  |  |  |
|  |  |  | Total number of service hours | 0.02 | NR | >0.05 |
|  |  |  | Step 5 |  |  |  |
|  |  |  | Time 2 number of optical aids | -0.08 | NR | <0.05 |
|  |  |  | Time 2 number of adaptive aids | 0.05 | NR | >0.05 |
| Rovner 2014 | PHQ-9 | Relative risk of incident depression | Randomization assignment | 0.5 | (0.3 to 1.0) | 0.037 |
|  |  |  | Stratum (20/70-20/100 vs <20/100) | 0.8 | (0.4 to 1.5) | 0.42 |
|  |  |  | Patient Health Questionnaire-9 | 1.1 | (0.9 to 1.2) | 0.53 |
|  |  |  | NEO-FFI neuroticism score | 1.0 | (1.0 to 1.1) | 0.071 |
|  |  |  | Medical Outcomes Study-6 | 1.1 | (1.0 to 1.2) | 0.014 |
| Rovner 2007 | HDRS | Odds ratio of depression diagnosis at 2 months | Treatment group | 2.3 | (1.1 to 5.1) | NR |
|  |  | Odds ratio of depression diagnosis at 2 months | Treatment group | 2.0 | (0.9 to 4.5) | NR |
|  |  |  | Activity loss | 2.6 | (1.2 to 5.5) | NR |
| Rees 2017 | PHQ-9 | PHQ-9 at 6 months | PST<sup>a</sup> | -3.7 | (-7.4 to -0.1) | 0.044 |
|  |  |  | PST<sup>b</sup> | -3.8 | (-7.3 to -0.2) | 0.037 |

Notes: <sup>a</sup>: Adjusted for baseline PST score; <sup>b</sup>: Adjusted for baseline PST score, age, gender, diabetes duration, diabetes medication, and visual acuity at baseline; SF-36: Medical Outcomes Study 36-Item Short Form Health Survey; TICS: Telephone Interview for Cognitive Status; NEO-FFI: Neuroticism, Extroversion, Openness Five Factor Inventory

**Table 5.**
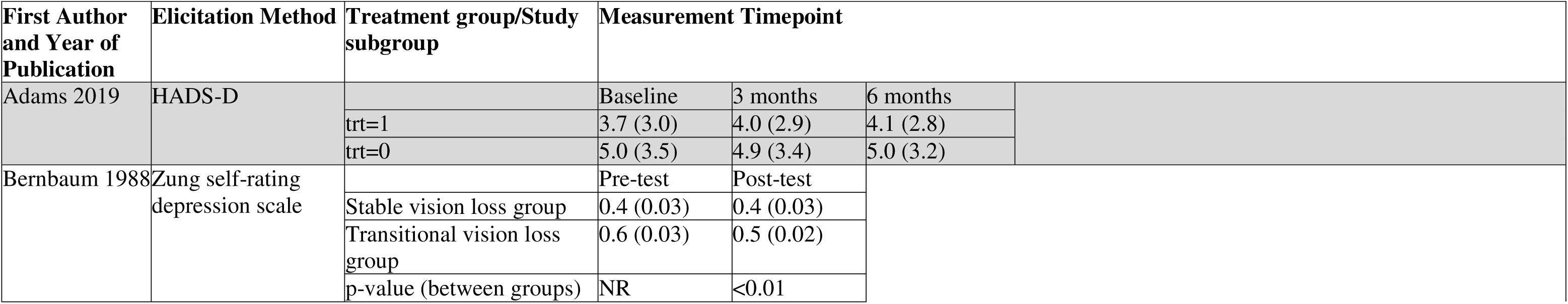
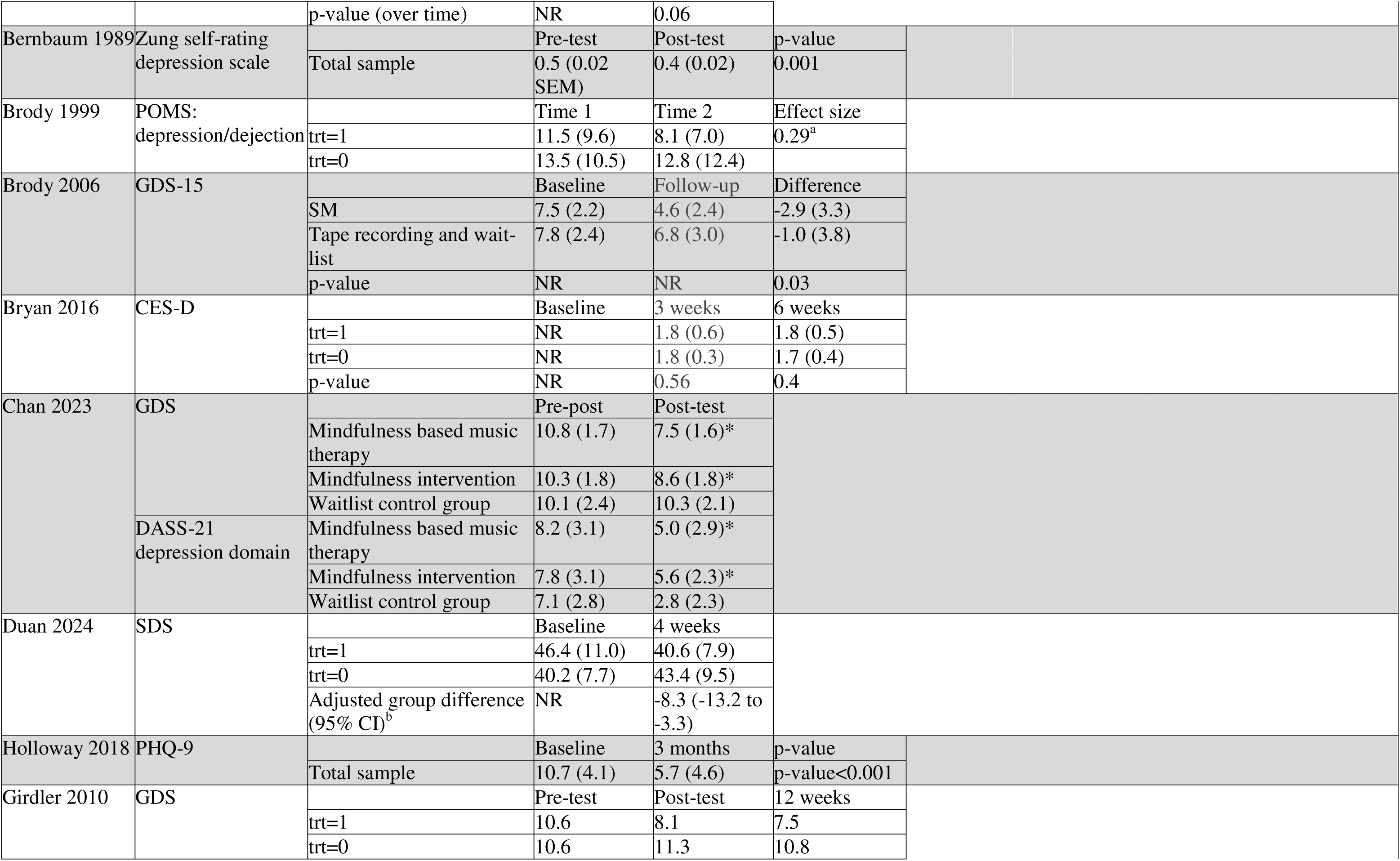
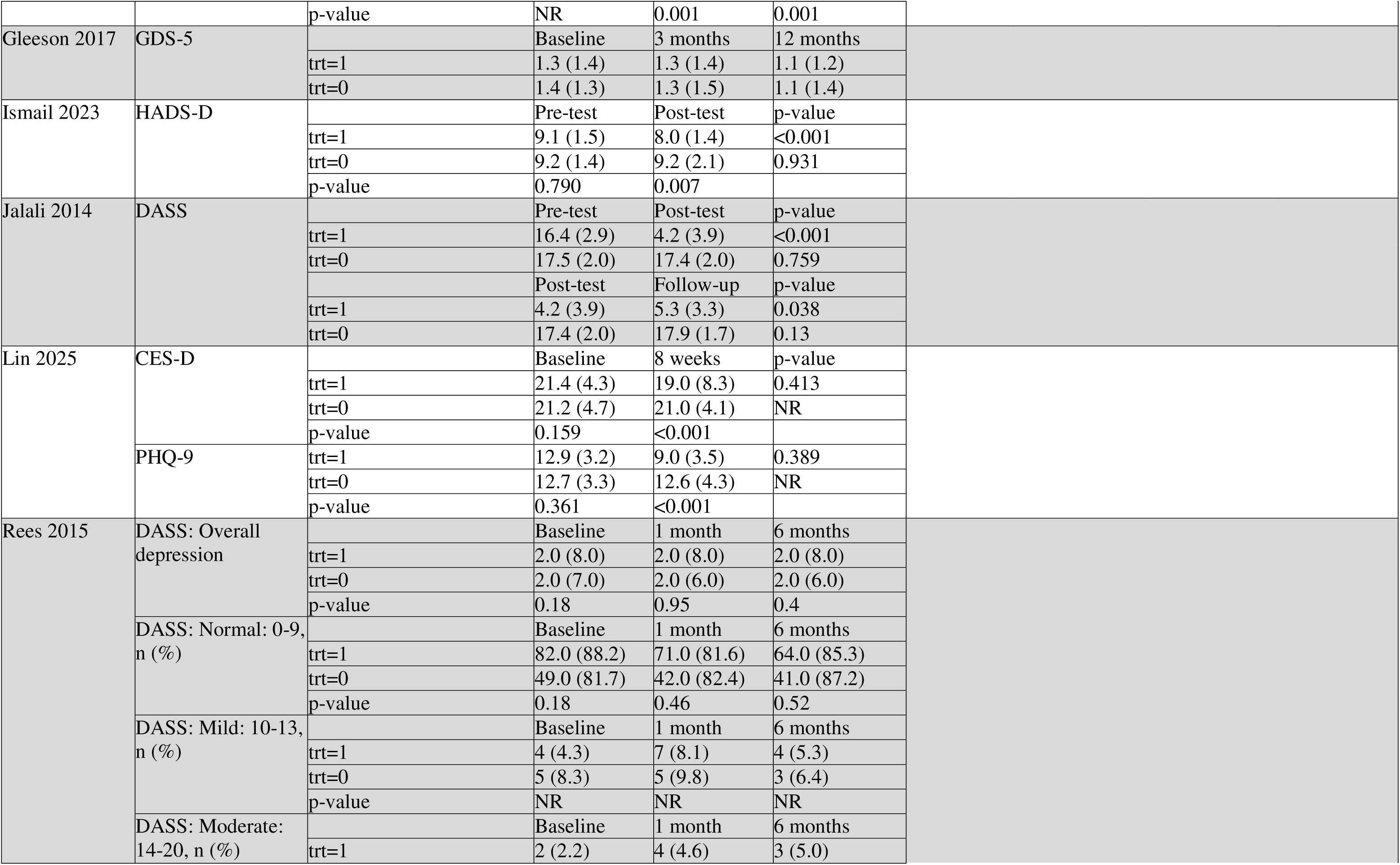
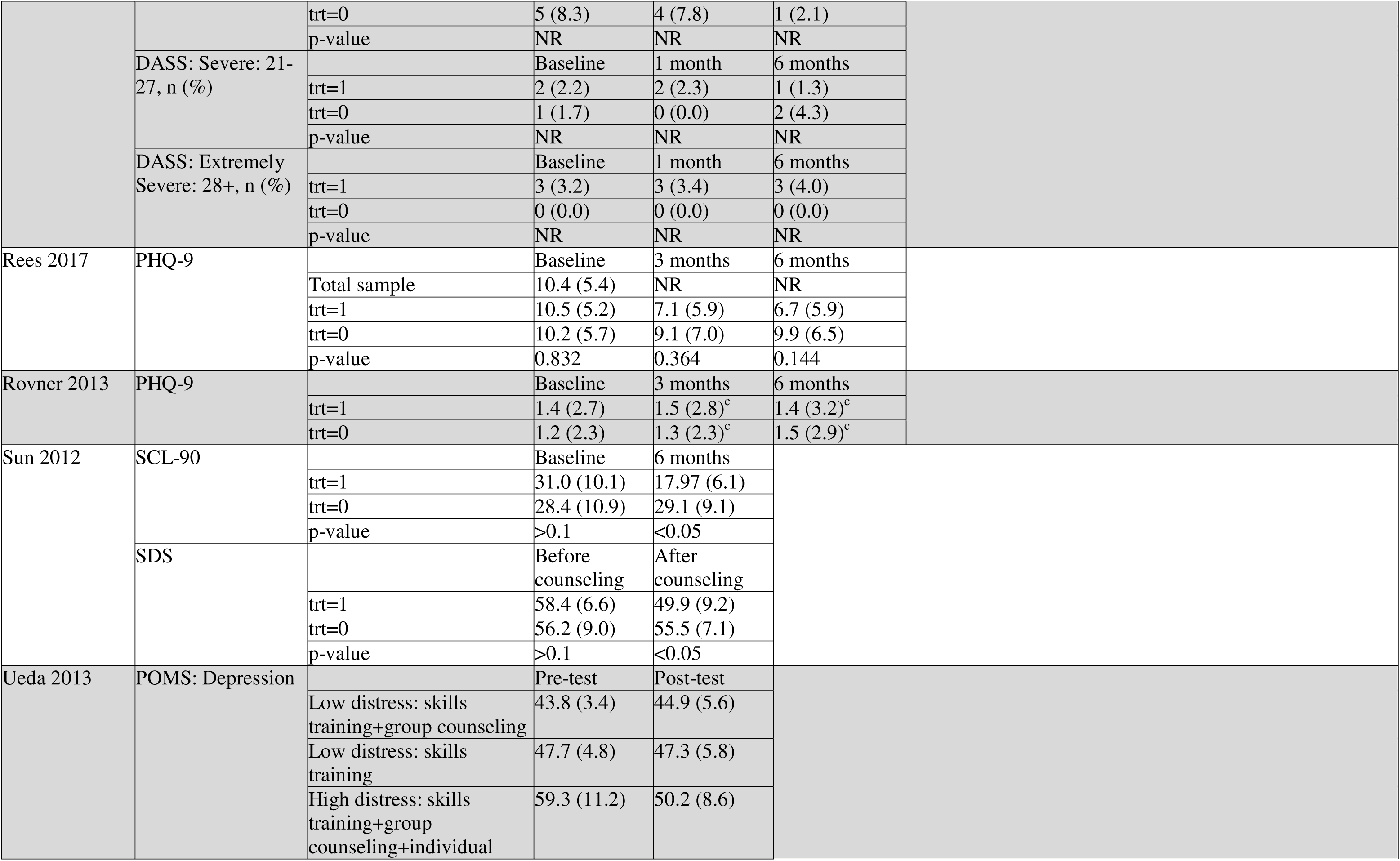
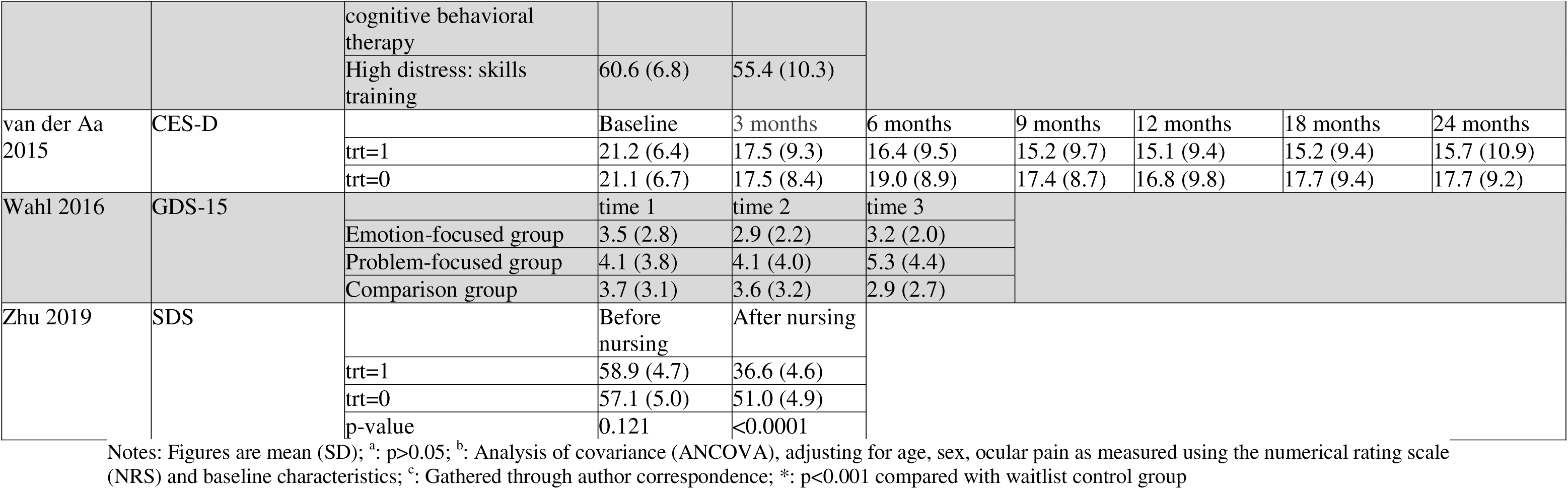
Depression Score at Each Measurement Timepoint.

The remaining 2 studies did not overlap in any dimension of the PICO framework other than the intervention and outcome. Holloway et al. (2018) measured the change in PHQ-9 score in a single group of eye disease patients administered PST (46). The results from this single-arm trial demonstrated a reduction in depression score from baseline (mean (SD)=10.7 (4.1)) to 3 months (mean (SD)=5.7 (4.6)) that reached statistical significance (p-value<0.001), however, there was significant attrition and data from only 18 of the 55 total enrolled participants contributed to data analysis (Table 5) (46). There was only 1 study that administered PST to diabetic retinopathy patients (64). In this study, the treatment group was administered PST and diabetes-specific educational brochures were given to participants in both groups (64). A regression model was plotted using Patient Health Questionnaire-9 (PHQ-9) score at 6 months as the outcome adjusted for treatment assignment, baseline score, age, gender, diabetes duration, diabetes medication, and visual acuity at baseline (64). Based on the regression output, the change from baseline to 3 months (p-value=0.03) and from baseline to 6 months (p-value=0.04) between treatment groups was significant (Table 6) (64).

**Table 6.** Change in Depression Score Across Each Measurement Timepoint.

| First Author and Year of Publication | Elicitation Method | Treatment group/Study subgroup | Measurement Timepoint |  |  |
| --- | --- | --- | --- | --- | --- |
| Adams 2019 | HADS-D |  | Baseline | Change from baseline to 3 months | Change from baseline to 6 months |
|  |  | trt=1 | 3.7 (3.0) | -0.1 (2.5) | 0.0 (2.3) |
|  |  | trt=0 | 5.0 (3.5) | -0.1 (2.2) | -0.1 (3.0) |
| Gleeson 2017 | GDS-5 |  | Baseline | Change from baseline to 3 months | Change from baseline to 12 months |
|  |  | trt=1 | 1.3 (1.4) | -0.1 (1.2) | -0.2 (1.1) |
|  |  | trt=0 | 1.4 (1.3) | -0.02 (1.1) | -0.2 (1.0) |
|  |  | trt=1 - trt=0 | NR | -0.05 (-0.45 to 0.34), p-value=0.78 | 0.01 (-0.34 to 0.37), p-value=0.93 |
| Kamga 2018 | PHQ-9 |  | Baseline | Change from baseline to 8 weeks |  |
|  |  | trt=1 | 9.5 (3.7) | -4.9 (4.2) |  |
|  |  | trt=0 | 9.5 (4.2) | -3.2 (4.6) |  |
|  |  | p-value | NR | 0.104<sup>a</sup>, 0.040<sup>b</sup> |  |
| Nollett 2016 | BDI |  | Baseline | Change from baseline to 3 months | Change from baseline to 6 months |
|  |  | PST | 19.0 (10.6) | -2.3 (5.8) | -2.7 (7.9) |
|  |  | Referral | 21.1 (7.6) | -1.1 (8.5) | -2.1 (7.6) |
|  |  | Waiting list | 20.3 (10.3) | -2.8 (10.4) | -1.1 (8.9) |
| Rees 2017 | PHQ-9 |  | Baseline | Change from baseline to 3 months | Change from baseline to 6 months |
|  |  | trt=1 | 10.5 (5.2) | -3.8 (3.0) | -4.3 (6.1) |
|  |  | trt=0 | 10.2 (5.7) | -1.1 (3.9) | -0.3 (4.6) |
|  |  | p-value | NR | 0.03 | 0.04 |
| Renieri 2013 | HADS-D |  | Baseline | After LVR | p-value |
|  |  | Participants | 6.2 (3.2) | 5.7 (3.1) | 0.235 |
|  |  | Dropouts | 5.7 (4.5) | NA |  |
|  |  | p-value | 0.568 | NR |  |
| Wahl 2006 | GDS-15 |  | Change from time 1 to time 2 | Change from time 1 to time 3 |  |
|  |  | Emotion-focused group | -0.6 | -0.04 |  |
|  |  | Problem-focused group | 0.2 | 1.4 |  |
|  |  | Comparison group | -0.06 | -0.3 |  |
|  |  | p-value<sup>e</sup> | 0.05 | 0.04 |  |

Notes: Figures are mean (SD); <sup>a</sup>: before adjusting for visual acuity; <sup>b</sup>: after adjusting for visual acuity; <sup>c</sup>: Analysis of variance (ANOVA); <sup>d</sup>: ANCOVA, adjusting for group differences at the baseline in the Problem Areas in Diabetes, Diabetes Knowledge test, or BDI; <sup>e</sup>: ANCOVA of intervention group effects on change from time 1 to time 2, controlled for time 1 levels

### 1.3.4 Self management

There were 4 studies that investigated the effectiveness of SM on depression and 2 of them administered SM to the same patient population. These studies administered SM to AMD patients (38,65). The first study compared SM to wait-list where participants in the latter group were administered SM after the conclusion of the first 6-week block (38). There was a decrease between baseline and follow-up in the depression/dejection domain of the Profile of Mood States (POMS), however, the effect size was not statistically significant (ES: 0.3, p>0.05) (Table 5) (38). The second study randomized AMD patients to 1 of 3 groups: SM, tape-recorded health education program, or wait-list (65). Patients assigned to the tape-recorded health education program were administered a series of health lectures that were equal in duration to the length of the SM program (65). There was no statistically significant difference in the change scores between participants in the tape-recorded health education program compared to the wait-list group, so the data from these two groups were combined to form a single control group (65). A mean (SD) change in Geriatric Depression Scale – 15 (GDS-15) score of -2.9 (3.3) and -1.0 (3.8) was observed among the SM and combined control group respectively (Table 5) (65). The difference in the change in depression score between the two groups was statistically significant (p-value=0.03) (65).

The remaining 2 studies compared SM to usual care on depression among vision impaired adults (47,62). There were key differences in the usual care interventions between the two studies. In the study conducted by Girdler et al. (2010), usual care included an initial interview with patients to jointly create a service plan, a visual assessment, and referral to the relevant service provider(s) (47). In contrast, in the study conducted by Rees et al. (2015), usual care constituted an initial assessment with a Low Vision Service (LVS) practitioner, optometric assessment and prescription of optical aids, and when requested, additional training was provided (62). Girdler et al. (2010) reported an average Geriatric Depression Scale (GDS) score of 10.6, 8.1, and 7.5 at pre-test, post-test and 3 months among the SM group (Table 5) (47). An average GDS score of 10.6, 11.3, and 10.8 was reported at the same measurement timepoints among the usual care group (47). A statistically significant difference between the treatment groups was found at both post-test (p-value=0.001) and 3 months (p-value=0.001) (47). In contrast, Rees et al. (2015) did not find a statistically significant difference in overall depression between SM and usual care at 1 month (p-value=0.95) and 6 months (p-value=0.4) using the Depression, Anxiety, Stress Scale (DASS) (Table 5) (62). The number of participants that fell within the normal range of depression symptoms was not statistically different between the two groups at 1 month (p-value=0.46) and 6 months (p-value=0.52) indicating that SM did not have a superior effect on depression symptoms compared to usual care (62).

### 1.3.5 Physical activity

Aside from the intervention and outcome, there was no overlap in terms of the PICO framework among the 3 studies that investigated the effectiveness of physical activity (36,54,59). In a single arm trial, an exercise protocol resulted in a borderline statistically significant improvement in SDS score over a 3 month study period among diabetic retinopathy patients (p-value=0.06) (Table 5) (36). In a prospective cohort study, high (compared to low) physical activity index scores at baseline were associated with fewer depression symptoms at follow-up as measured using the Hospital Anxiety Depression Scale - Depression following adjustment for potential confounders (Table 7) (54). These associations were only statistically significant among the self-reported no visual impairment (SRNI) and among women in the self-reported visual impairment (SRVI) group (54). In the RCT conducted by Adams et al. (2019), visually impaired adults were randomized to a exercise program compared to a usual care arm where no intervention was administered (59). The 3 month exercise program consisted of weekly sessions that lasted up to an hour (59). Participants in the treatment arm were also instructed to exercise independently at home for up to 2 hours per week (59). The mean (SD) HADS-D scores at baseline, 3 months, and 6 months were 3.7 (3.0), 4.0 (2.9), 4.1 (2.8) respectively for the treatment group and 5.0 (3.5), 4.9 (3.4), and 5.0 (3.2) respectively for the usual care group (59). The mean (SD) change from baseline to 3 months and from baseline to 6 months was -0.1 (2.5) and 0.0 (2.3) respectively for the treatment group and -0.1 (2.2) and -0.1 (3.0) respectively for the usual care group (59). Although the authors did not calculate statistical significance, the median change from baseline to 3 and from baseline to 6 months was 0 suggesting a lack of meaningful differences within groups across time (59).

**Table 7.**
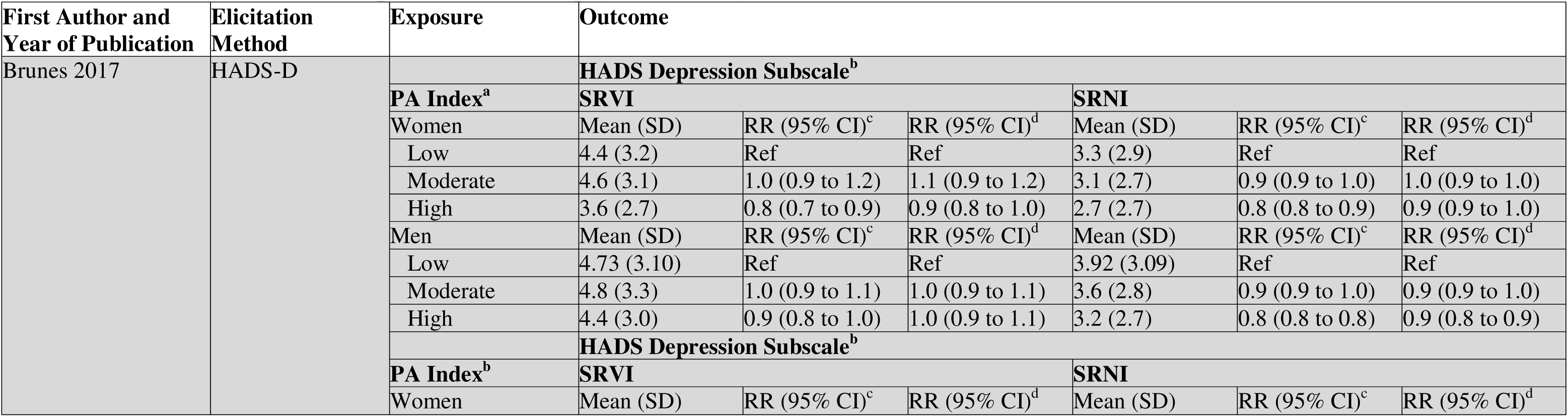
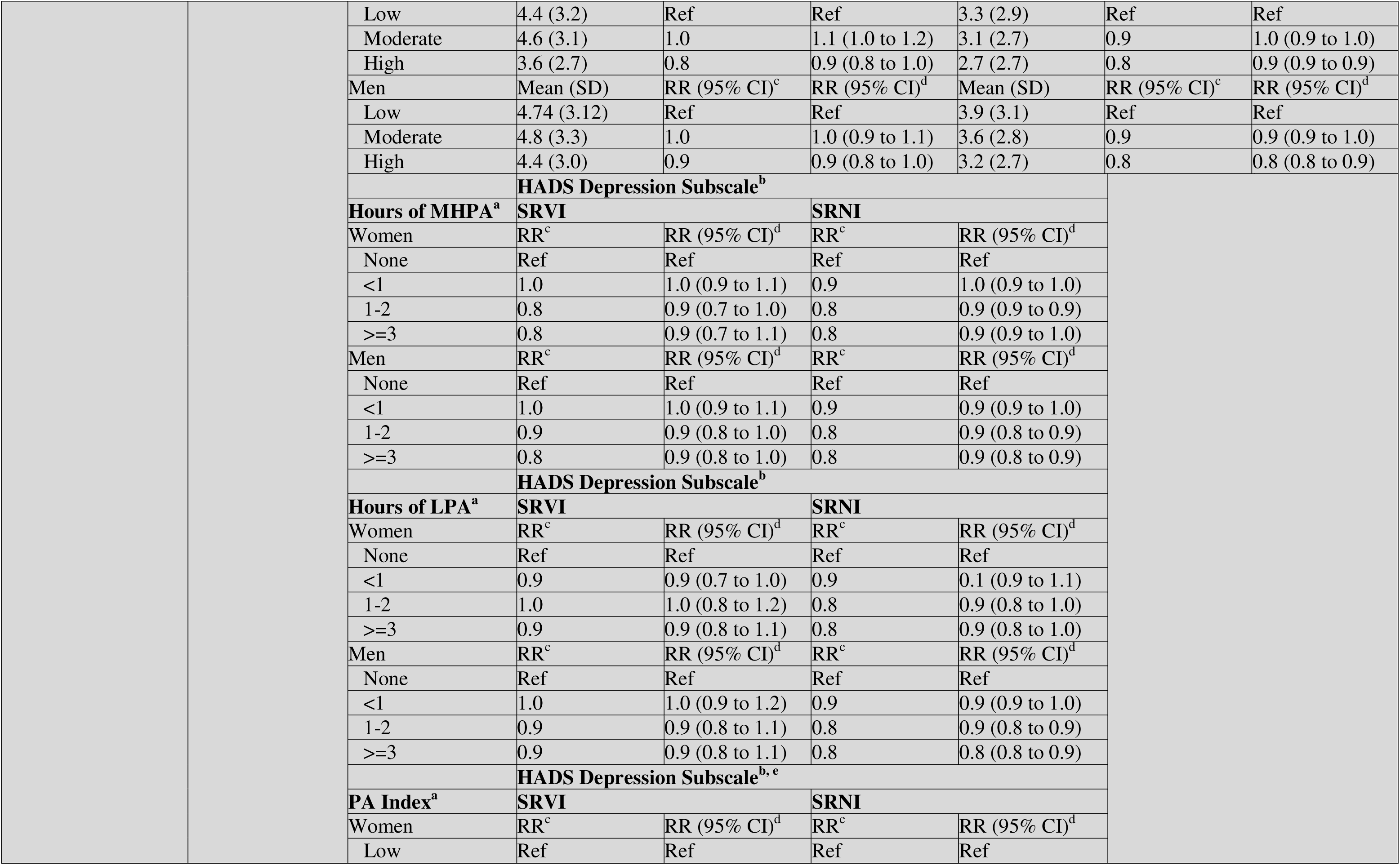
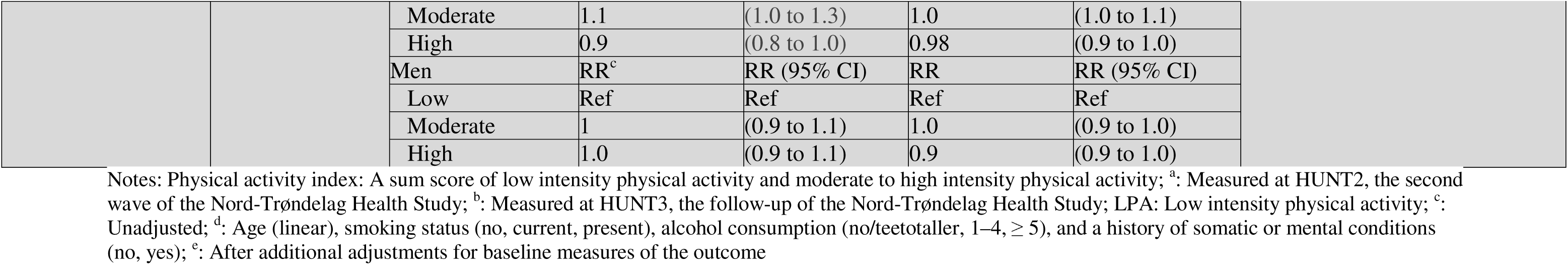
Risk ratio for depression based on physical activity index^a^.

### 1.3.6 Risk of Bias

Given the nature of non-pharmacological interventions, none of the patients or healthcare providers in the randomized control trials were blinded (Table 8) (35,38,40,42–45,47,49–51,55,56,58–65). Although outcome assessors in Girdler et al. (2010) were masked to group allocation, participants often revealed their allocation status during follow-up interviews (47). In Wahl et al. (2006), participants in the emotion- or problem-focused group were randomly assigned to their respective treatment group but the participants in the comparison group were not due to limited resources and were instead, selected from an outpatient list (49). The data analysts were not blinded in the studies conducted by Rovner et al. (2007) and Rovner et al. (2014) (35,43). In addition, only the outcome assessors were blinded in the study conducted by Ismail et al. (2023) (60). A total of 3 randomized control trials suffered from substantial attrition bias (>20%) (40,58,64). The cohort studies were at low risk of bias (Table 9).

**Table 8.** Tool to Assess Risk of Bias in Randomized Controlled Trials.

| First Author and Year of Publication | 1 Was the allocation sequence adequately generated? | 2 Was the allocation sequence adequately concealed? | 3.a. Were patients blinded? | 3.b. Were healthcare providers blinded? | 3.c. Were data collectors blinded? | 3.d. Were outcome assessors blinded? | 3.e. Were data analysts blinded? | 4 Was loss to follow-up (missing outcome data) infrequent? | 5 Are reports of the study free of selective outcome reporting? | 6 Was the study apparently free of other problems that could put it at a risk of bias? |
| --- | --- | --- | --- | --- | --- | --- | --- | --- | --- | --- |
| Adams 2019 | Y | Y | N | N | PN | PN | PY | Y | Y | PY |
| Chan 2023 | Y | Y | N | N | Y | PY | Y | Y | Y | PY |
| van der Aa 2015 | Y | Y | N | N | Y | Y | PY | N | PY | PY |
| Gleeson 2017 | Y | Y | N | N | Y | Y | PY | Y | Y | PY |
| Kamga 2018 | Y | PY | N | N | Y | PY | PY | PY | Y | PY |
| Nollett 2016 | Y | Y | N | N | Y | Y | PY | Y | Y | PY |
| Rees 2017 | Y | Y | N | N | Y | Y | PY | N | Y | PY |
| Rovner 2007 | Y | Y | N | N | Y | Y | N | Y | Y | Y |
| Rovner 2014 | Y | Y | N | N | Y | Y | N | PN | Y | PY |
| Sun 2012 | PY | PY | N | N | PY | PY | PY | PY | PN | Y |
| Zhu 2019 | Y | PY | N | N | PY | PY | PY | PY | PN | Y |
| Brody 1999 | PY | PY | N | N | PY | PY | PY | Y | PY | PY |
| Brody 2006 | Y | Y | N | N | Y | Y | PY | Y | PY | PY |
| Bryan 2016 | Y | PY | N | N | PY | PY | PY | N | PN | PY |
| Girdler 2010 | Y | PY | N | N | PY | N | PY | Y | PY | PY |
| Ismail 2023 | PY | Y | N | N | N | Y | N | Y | Y | PY |
| Jalali 2014 | PY | PY | N | N | PY | PY | PY | Y | PN | PY |
| Rees 2015 | Y | Y | N | N | Y | Y | PY | PN | PY | PY |
| Rovner 2013 | Y | Y | N | N | Y | Y | N | PN | PN | PY |
| Wahl 2006 | PY | PY | N | N | PY | PY | PY | PN | PN | N |

Notes: Y=Yes; PY=Probably yes; PN=Probably no; N=No

**Table 9.** Tool to Assess Risk of Bias in Cohort Studies.

| First Author and Year of Publication | 1. Was selection of exposed and non-exposed | 2. Can we be confident in the assessment of | 3. Can we be confident that the outcome of | 4. Did the study match exposed and unexposed | 5. Can we be confident in the assessment of the | 6. Can we be confident in the assessment of | 7. Was the follow up of cohorts adequate? | 8. Were cointerventions similar between |
| --- | --- | --- | --- | --- | --- | --- | --- | --- |

|  | cohorts drawn from the same population? | exposure? | interest was not present at the start of the study? | for all variables that are associated with the outcome of interest or did the statistical analysis adjust for these prognostic variables? | presence or absence of prognostic factors? | outcome? |  | groups? |
| --- | --- | --- | --- | --- | --- | --- | --- | --- |
| Brunes 2017 | Y | Y | PY | Y | Y | Y | Y | Y |
| Horowitz 2006 | Y | Y | Y | Y | Y | Y | Y | PY |
| Lin 2025 | Y | Y | Y | N | Y | Y | PY | PY |

Of the non-randomized studies, there 3 studies that did not control for confounding (Table 10) (36,46,57). A total of 2 studies suffered from bias due to missing data (34,48). Renieri et al. (2013) suffered from substantial attrition bias whereas Trozzolino et al. (2003) were not able to conduct blood tests to assess diabetes control among participants in the comparison arm as a result of limited resources (34,48).

**Table 10.** Risk of Bias in Non-Randomized Studies - of Interventions (ROBINS-I)

| First Author and Year of Publication | Bias due to confounding | Bias due to selection of participants | Bias in classification of interventions | Bias due to deviations from intended interventions | Bias due to missing data | Bias in measurement of outcomes | Bias in selection of the reported result | Overall bias |
| --- | --- | --- | --- | --- | --- | --- | --- | --- |
| Bernbaum 1988 | Serious | Low | Low | Low | Low | Low | Low | Moderate |
| Bernbaum 1989 | Moderate | Low | Low | Low | Low | Low | Low | Moderate |
| Duan 2024 | Moderate | Low | Low | Low | Low | Low | Low | Low |
| Holloway 2018 | Serious | Low | Low | Low | Moderate | Low | Low | Moderate |
| Renieri 2013 | Low | Low | Low | Low | Serious | Low | Low | Moderate |
| Trozzolino 2003 | Low | Low | Low | Low | Serious | Low | Low | Moderate |
| Ueda 2013 | Serious | Low | Low | Low | Low | Low | Low | Moderate |

## 1.4 Discussion

Based on the results of the qualitative analyses from the current review, there were conflicting findings regarding the effectiveness of PST among eye disease patients. To the extent that PST is effective in improving depression symptoms among older adults with eye disease, its effectiveness may be attributed to its role in preventing loss of a valued activity by providing them with skills to form alternative solutions in spite of their visual impairment (43). This finding is in accordance with existing literature demonstrating that activity restriction may negatively impact psychological health (66). Interestingly, PST demonstrated a statistically significant effect on depression compared to usual care at 2 months but not at 6 months (43). The point estimate of the OR for depression at 6 months was below 1 but the confidence intervals crossed the null effect (43). This indicates that PST may have a maintenance effect on depression but that there is wide variability in this effect (43). This suggests that booster treatments may be necessary to sustain the positive impact of PST on depression. It is worth noting that psychosocial interventions such as PST are most effective in addressing subsyndromal and mild depression (67). Considering this, the variation in the effectiveness of PST may be attributed, in part, to varying depression severity between studies.

It is plausible that the skills gained from PST decay over time, especially among eye disease patients that experience normal age-related cognitive decline (68) in addition to any impairment in cognitive functioning resulting from visual impairment (69). Indeed, existing literature points to the necessity of booster treatment for maintaining the long-term effectiveness of PST. In an RCT assigning older adults with subsyndromal depression to a PST and brief behavioral treatment for insomnia versus usual care, the effectiveness of the treatment was maintained at 12 months after a booster treatment at 7 and 10 months (70). In a 3-arm RCT, homebound older adults were randomized to tele-PST, in-person PST, or telephone support calls (71). In this study, 6 sessions were administered for each treatment arm with 6 monthly booster sessions conducted by telephone (71). There was a reduction in depression among the tele-PST and in-person PST groups compared to the telephone support call group at 3 months and this effect was maintained at 6 months (71).

There were inconclusive results regarding the effectiveness of SM most likely as a result of the poor methodological quality of included studies. One study compared SM to wait-list (38) and another compared SM to tape recorded health education program or wait-list (65) among AMD patients. Both studies suffered from small sample sizes (38,65) and one study conducted by Brody et al. (2006) suffered from co-intervention bias resulting from concomitant antidepressant use among patients in both experimental and control group (65). In this study, all 3 of the participants in the experimental group and 4 of the 7 participants in the control group demonstrated a clinically meaningful improvement in depression symptoms (65). The two included studies that examined SM versus usual care revealed contradicting results as well (47,62). The RCT conducted by Rees et al. (2015) revealed no statistically significant difference in depression score between the SM and usual care group at 1 month (p-value=0.95) and 6 months (p-value=0.4) (62). This may be attributed, in part, to inconsistencies in how SM was administered to participants. Although the healthcare practitioners were experienced, formal training on how to conduct SM was not provided by the study investigators (62). The authors also reported that the healthcare practitioners likely modified the structure of the program to suit the needs of different participants (62). Girdler et al. (2010) conducted a methodologically rigorous RCT and demonstrated a statistically significant difference in mean GDS score at post-test (p-value=0.001) and 12 weeks (p-value=0.001) (47). Notably, depression scores worsened in the usual care arm providing further evidence for the need to conduct more extensive research into viable mental health treatments for eye disease patients (47).

The mixed results regarding the relationship between physical activity and depression among eye disease patients are likely attributed to the variation in the amount and type of exercise that was investigated across included studies. Adams et al. (2019) randomized visually impaired patients to a combination of aerobic, resistance, and mobility exercises for 110 minutes per week targeted towards fall prevention which resulted in minimal changes in HADS-D score over time (59). Although the duration of the exercise sessions was not documented, Bernbaum et al. (1988) administered 3 mobility and aerobic exercise sessions per week to participants which was associated with a borderline statistically significant improvement in depression score over time (p-value=0.06) among diabetic retinopathy patients (36). Brunes et al. (2017) demonstrated that high baseline physical activity was associated with fewer depression symptoms at follow-up among both SRNI and SRVI participants, but this relationship only reached statistical significance among the SRNI group and women in the SRVI group (54). Indeed, scoring high on the physical activity index is indicative of meeting the WHO physical activity recommendation which is necessary to maintain or improve functioning For context, the WHO recommends that adults engage in 150-300 minutes of moderate physical activity per week (72).

The notion that the heterogeneity observed in the current study can be explained by variability in the exercises that participants underwent is supported by evidence from previous research demonstrating a dose-response relationship between exercise and depression. Interestingly, a minimum effective dose of approximately 97.5 minutes of moderate physical activity per week was observed in a network meta-analysis (73) which is lower than the WHO recommendation. This is encouraging and suggests that older adults may be able to reap the benefits of physical activity at lower doses than anticipated. Based on results from the same review, the dose-response relationship between physical activity and depression also varies based on the modality of exercise (72). Although various modalities of physical activity improved depression among older adults, resistance training (exercises that were performed with the objective of increasing muscle strength and endurance) and mind-body exercises (interventions that seek to integrate the mind, body, and spirit such as yoga, tai chi, and qi gong) in particular had the most substantial impact (73). Resistance exercise may indirectly ameliorate an individual’s quality of life and reduce depressive symptoms by improving their body composition and metabolic health (74) to an extent that can not be achieved by aerobic exercise or mixed modalities. In contrast, mind-body exercises seem to have a positive impact on depression among older adults by maintaining a balanced mental state (75).

### 1.4.1 Limitations

Caution must be exercised when interpreting the results of the current systematic review. Although the results of the data extraction were reviewed by all authors, data were only extracted by a single author (J.S.). Based on the risk of assessment of included studies, none of the RCTs blinded patients or healthcare practitioners to the respective intervention administered (35,38,40,42–45,47,49–51,55,56,58–62,64,65). In addition, the majority of non-randomized studies failed to adjust for potential confounders (36,37,46,57). The findings from the current review are also limited by the heterogeneity in eye diseases, interventions, and comparators among the included studies which makes comparisons between studies difficult to make. Lastly, the majority of participants in the included studies suffered from mild to moderate depression at baseline. Therefore, the results of this study can not be generalized to severe depression.

### 1.4.2 Future Directions

In the future, methodologically rigorous studies should be conducted to investigate the necessity for booster treatments to maintain the positive effects of PST. Subgroup analyses should moreover be conducted to determine whether depression type (i.e., MMD, persistent depressive disorder (PDD), postpartum depressive disorder (PPD), etc.) and severity influences the effectiveness of non-pharmacological treatment effectiveness. RCTs controlling for the amount of exercise per week should be conducted to assess the effectiveness of physical activity in treating depression among visually impaired older adults. Lastly, interventions should be tailored to accommodate the functional impairment brought about by eye disease and other comorbid conditions among this population.

## 1.5 Conclusions

Physical activity may be a viable intervention to treat depression among visually impaired older adults, however, randomized data accounting for physical activity per week are necessary before coming to a firm conclusion. Although some studies have demonstrated a short benefit for PST, further research is necessary to determine the long-term effectiveness of this treatment as well as the necessity for booster treatment. Finally, future studies should stratify results by depression type and severity to determine if this impacts treatment effectiveness.

## Data Availability

All relevant data are within the manuscript

## Data availability statement

All relevant data are within the paper

## 1.1 Appendix

## Appendix 1. Search Strategy

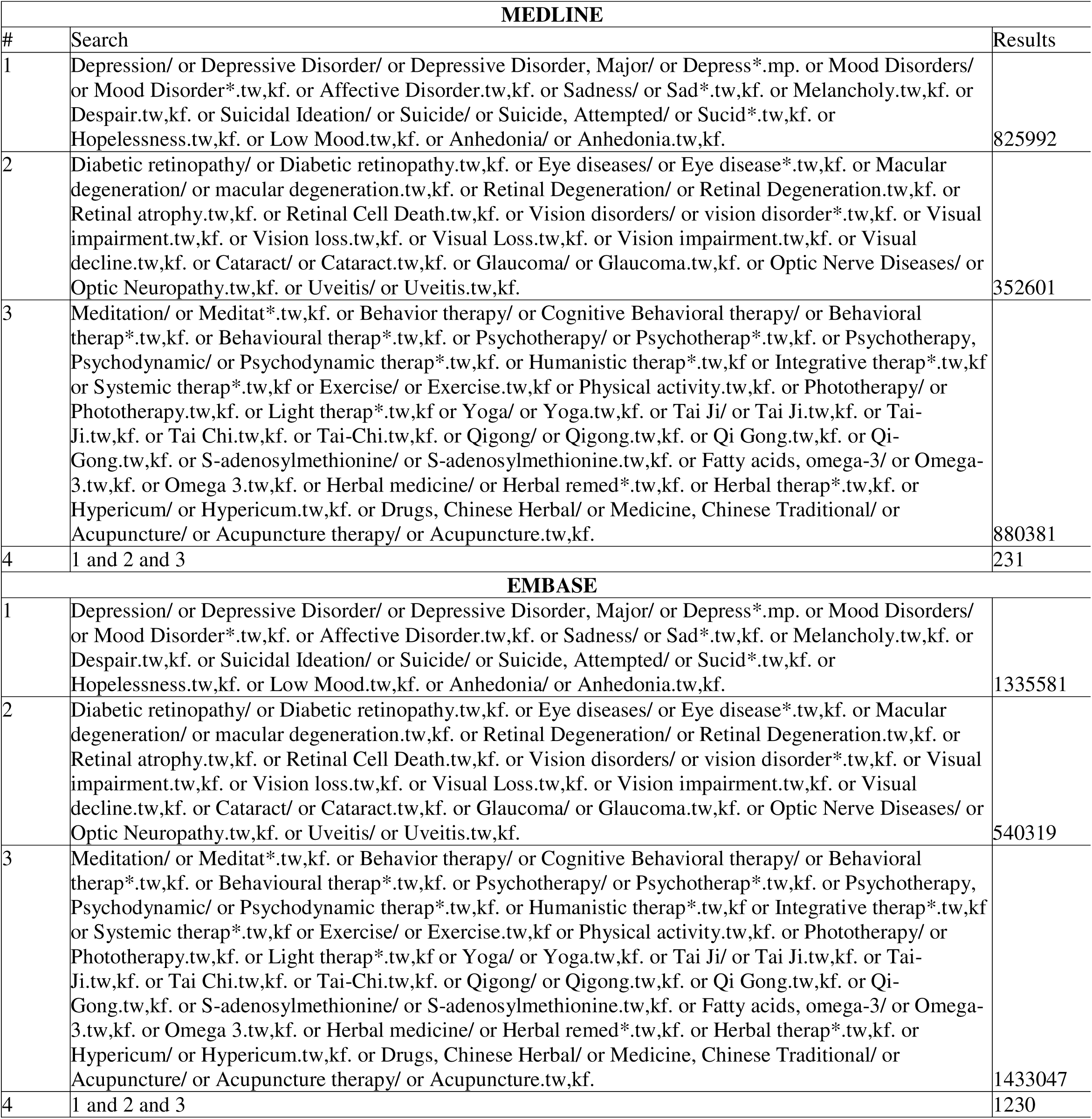
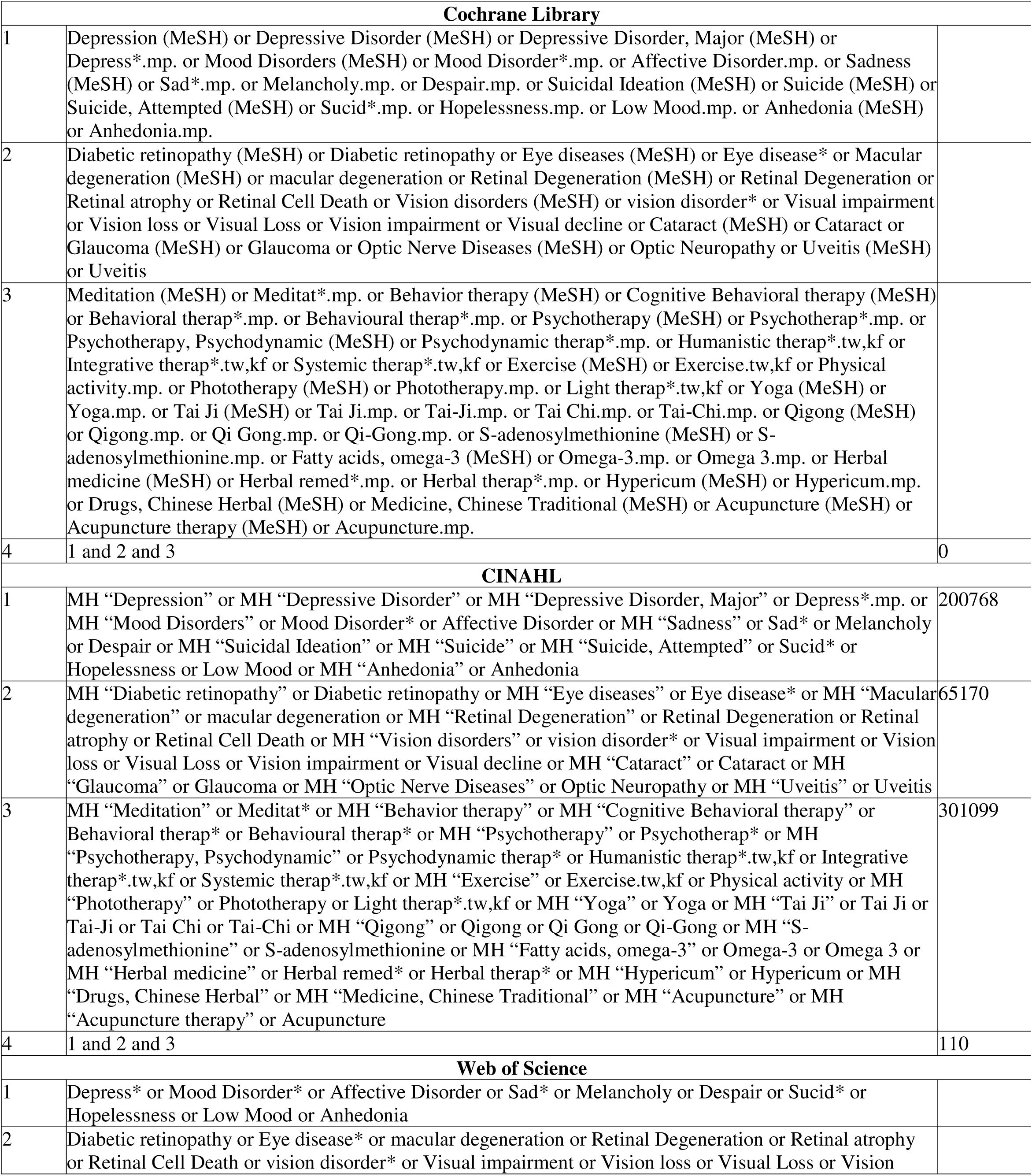
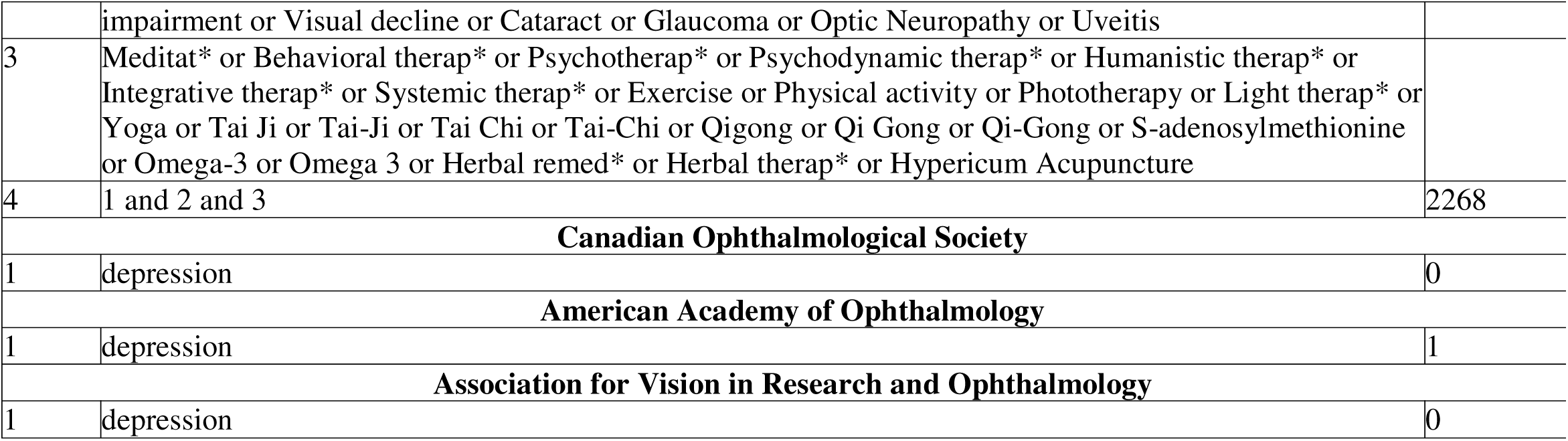

| MEDLINE |  |  |
| --- | --- | --- |
| # | Search | Results |
| 1 | Depression/ or Depressive Disorder/ or Depressive Disorder, Major/ or Depress*.mp. or Mood Disorders/ or Mood Disorder*.tw,kf. or Affective Disorder.tw,kf. or Sadness/ or Sad*.tw,kf. or Melancholy.tw,kf. or Despair.tw,kf. or Suicidal Ideation/ or Suicide/ or Suicide, Attempted/ or Sucid*.tw,kf. or Hopelessness.tw,kf. or Low Mood.tw,kf. or Anhedonia/ or Anhedonia.tw,kf. | 825992 |
| 2 | Diabetic retinopathy/ or Diabetic retinopathy.tw,kf. or Eye diseases/ or Eye disease*.tw,kf. or Macular degeneration/ or macular degeneration.tw,kf. or Retinal Degeneration/ or Retinal Degeneration.tw,kf. or Retinal atrophy.tw,kf. or Retinal Cell Death.tw,kf. or Vision disorders/ or vision disorder*.tw,kf. or Visual impairment.tw,kf. or Vision loss.tw,kf. or Visual Loss.tw,kf. or Vision impairment.tw,kf. or Visual decline.tw,kf. or Cataract/ or Cataract.tw,kf. or Glaucoma/ or Glaucoma.tw,kf. or Optic Nerve Diseases/ or Optic Neuropathy.tw,kf. or Uveitis/ or Uveitis.tw,kf. | 352601 |
| 3 | Meditation/ or Meditat*.tw,kf. or Behavior therapy/ or Cognitive Behavioral therapy/ or Behavioral therap*.tw,kf. or Behavioural therap*.tw,kf. or Psychotherapy/ or Psychotherap*.tw,kf. or Psychotherapy, Psychodynamic/ or Psychodynamic therap*.tw,kf. or Humanistic therap*.tw,kf. or Integrative therap*.tw,kf. or Systemic therap*.tw,kf. or Exercise/ or Exercise.tw,kf. or Physical activity.tw,kf. or Phototherapy/ or Phototherapy.tw,kf. or Light therap*.tw,kf. or Yoga/ or Yoga.tw,kf. or Tai Ji/ or Tai Ji.tw,kf. or Tai-Ji.tw,kf. or Tai Chi.tw,kf. or Tai-Chi.tw,kf. or Qigong/ or Qigong.tw,kf. or Qi Gong.tw,kf. or Qi-Gong.tw,kf. or S-adenosylmethionine/ or S-adenosylmethionine.tw,kf. or Fatty acids, omega-3/ or Omega-3.tw,kf. or Omega 3.tw,kf. or Herbal medicine/ or Herbal remed*.tw,kf. or Herbal therap*.tw,kf. or Hypericum/ or Hypericum.tw,kf. or Drugs, Chinese Herbal/ or Medicine, Chinese Traditional/ or Acupuncture/ or Acupuncture therapy/ or Acupuncture.tw,kf. | 880381 |
| 4 | 1 and 2 and 3 | 231 |
| EMBASE |  |  |
| 1 | Depression/ or Depressive Disorder/ or Depressive Disorder, Major/ or Depress*.mp. or Mood Disorders/ or Mood Disorder*.tw,kf. or Affective Disorder.tw,kf. or Sadness/ or Sad*.tw,kf. or Melancholy.tw,kf. or Despair.tw,kf. or Suicidal Ideation/ or Suicide/ or Suicide, Attempted/ or Sucid*.tw,kf. or Hopelessness.tw,kf. or Low Mood.tw,kf. or Anhedonia/ or Anhedonia.tw,kf. | 1335581 |
| 2 | Diabetic retinopathy/ or Diabetic retinopathy.tw,kf. or Eye diseases/ or Eye disease*.tw,kf. or Macular degeneration/ or macular degeneration.tw,kf. or Retinal Degeneration/ or Retinal Degeneration.tw,kf. or Retinal atrophy.tw,kf. or Retinal Cell Death.tw,kf. or Vision disorders/ or vision disorder*.tw,kf. or Visual impairment.tw,kf. or Vision loss.tw,kf. or Visual Loss.tw,kf. or Vision impairment.tw,kf. or Visual decline.tw,kf. or Cataract/ or Cataract.tw,kf. or Glaucoma/ or Glaucoma.tw,kf. or Optic Nerve Diseases/ or Optic Neuropathy.tw,kf. or Uveitis/ or Uveitis.tw,kf. | 540319 |
| 3 | Meditation/ or Meditat*.tw,kf. or Behavior therapy/ or Cognitive Behavioral therapy/ or Behavioral therap*.tw,kf. or Behavioural therap*.tw,kf. or Psychotherapy/ or Psychotherap*.tw,kf. or Psychotherapy, Psychodynamic/ or Psychodynamic therap*.tw,kf. or Humanistic therap*.tw,kf. or Integrative therap*.tw,kf. or Systemic therap*.tw,kf. or Exercise/ or Exercise.tw,kf. or Physical activity.tw,kf. or Phototherapy/ or Phototherapy.tw,kf. or Light therap*.tw,kf. or Yoga/ or Yoga.tw,kf. or Tai Ji/ or Tai Ji.tw,kf. or Tai-Ji.tw,kf. or Tai Chi.tw,kf. or Tai-Chi.tw,kf. or Qigong/ or Qigong.tw,kf. or Qi Gong.tw,kf. or Qi-Gong.tw,kf. or S-adenosylmethionine/ or S-adenosylmethionine.tw,kf. or Fatty acids, omega-3/ or Omega-3.tw,kf. or Omega 3.tw,kf. or Herbal medicine/ or Herbal remed*.tw,kf. or Herbal therap*.tw,kf. or Hypericum/ or Hypericum.tw,kf. or Drugs, Chinese Herbal/ or Medicine, Chinese Traditional/ or Acupuncture/ or Acupuncture therapy/ or Acupuncture.tw,kf. | 1433047 |
| 4 | 1 and 2 and 3 | 1230 |
| Cochrane Library |  |  |
| 1 | Depression (MeSH) or Depressive Disorder (MeSH) or Depressive Disorder, Major (MeSH) or Depress*.mp. or Mood Disorders (MeSH) or Mood Disorder*.mp. or Affective Disorder.mp. or Sadness (MeSH) or Sad*.mp. or Melancholy.mp. or Despair.mp. or Suicidal Ideation (MeSH) or Suicide (MeSH) or Suicide, Attempted (MeSH) or Sucid*.mp. or Hopelessness.mp. or Low Mood.mp. or Anhedonia (MeSH) or Anhedonia.mp. |  |
| 2 | Diabetic retinopathy (MeSH) or Diabetic retinopathy or Eye diseases (MeSH) or Eye disease* or Macular degeneration (MeSH) or macular degeneration or Retinal Degeneration (MeSH) or Retinal Degeneration or Retinal atrophy or Retinal Cell Death or Vision disorders (MeSH) or vision disorder* or Visual impairment or Vision loss or Visual Loss or Vision impairment or Visual decline or Cataract (MeSH) or Cataract or Glaucoma (MeSH) or Glaucoma or Optic Nerve Diseases (MeSH) or Optic Neuropathy or Uveitis (MeSH) or Uveitis |  |
| 3 | Meditation (MeSH) or Meditat*.mp. or Behavior therapy (MeSH) or Cognitive Behavioral therapy (MeSH) or Behavioral therap*.mp. or Behavioural therap*.mp. or Psychotherapy (MeSH) or Psychotherap*.mp. or Psychotherapy, Psychodynamic (MeSH) or Psychodynamic therap*.mp. or Humanistic therap*.tw,kf or Integrative therap*.tw,kf or Systemic therap*.tw,kf or Exercise (MeSH) or Exercise.tw,kf or Physical activity.mp. or Phototherapy (MeSH) or Phototherapy.mp. or Light therap*.tw,kf or Yoga (MeSH) or Yoga.mp. or Tai Ji (MeSH) or Tai Ji.mp. or Tai-Ji.mp. or Tai Chi.mp. or Tai-Chi.mp. or Qigong (MeSH) or Qigong.mp. or Qi Gong.mp. or Qi-Gong.mp. or S-adenosylmethionine (MeSH) or S-adenosylmethionine.mp. or Fatty acids, omega-3 (MeSH) or Omega-3.mp. or Omega 3.mp. or Herbal medicine (MeSH) or Herbal remed*.mp. or Herbal therap*.mp. or Hypericum (MeSH) or Hypericum.mp. or Drugs, Chinese Herbal (MeSH) or Medicine, Chinese Traditional (MeSH) or Acupuncture (MeSH) or Acupuncture therapy (MeSH) or Acupuncture.mp. |  |
| 4 | 1 and 2 and 3 | 0 |
| CINAHL |  |  |
| 1 | MH "Depression" or MH "Depressive Disorder" or MH "Depressive Disorder, Major" or Depress*.mp. or MH "Mood Disorders" or Mood Disorder* or Affective Disorder or MH "Sadness" or Sad* or Melancholy or Despair or MH "Suicidal Ideation" or MH "Suicide" or MH "Suicide, Attempted" or Sucid* or Hopelessness or Low Mood or MH "Anhedonia" or Anhedonia | 200768 |
| 2 | MH "Diabetic retinopathy" or Diabetic retinopathy or MH "Eye diseases" or Eye disease* or MH "Macular degeneration" or macular degeneration or MH "Retinal Degeneration" or Retinal Degeneration or Retinal atrophy or Retinal Cell Death or MH "Vision disorders" or vision disorder* or Visual impairment or Vision loss or Visual Loss or Vision impairment or Visual decline or MH "Cataract" or Cataract or MH "Glaucoma" or Glaucoma or MH "Optic Nerve Diseases" or Optic Neuropathy or MH "Uveitis" or Uveitis | 65170 |
| 3 | MH "Meditation" or Meditat* or MH "Behavior therapy" or MH "Cognitive Behavioral therapy" or Behavioral therap* or Behavioural therap* or MH "Psychotherapy" or Psychotherap* or MH "Psychotherapy, Psychodynamic" or Psychodynamic therap* or Humanistic therap*.tw,kf or Integrative therap*.tw,kf or Systemic therap*.tw,kf or MH "Exercise" or Exercise.tw,kf or Physical activity or MH "Phototherapy" or Phototherapy or Light therap*.tw,kf or MH "Yoga" or Yoga or MH "Tai Ji" or Tai Ji or Tai-Ji or Tai Chi or Tai-Chi or MH "Qigong" or Qigong or Qi Gong or Qi-Gong or MH "S-adenosylmethionine" or S-adenosylmethionine or MH "Fatty acids, omega-3" or Omega-3 or Omega 3 or MH "Herbal medicine" or Herbal remed* or Herbal therap* or MH "Hypericum" or Hypericum or MH "Drugs, Chinese Herbal" or MH "Medicine, Chinese Traditional" or MH "Acupuncture" or MH "Acupuncture therapy" or Acupuncture | 301099 |
| 4 | 1 and 2 and 3 | 110 |
| Web of Science |  |  |
| 1 | Depress* or Mood Disorder* or Affective Disorder or Sad* or Melancholy or Despair or Sucid* or Hopelessness or Low Mood or Anhedonia |  |
| 2 | Diabetic retinopathy or Eye disease* or macular degeneration or Retinal Degeneration or Retinal atrophy or Retinal Cell Death or vision disorder* or Visual impairment or Vision loss or Visual Loss or Vision |  |
|  | impairment or Visual decline or Cataract or Glaucoma or Optic Neuropathy or Uveitis |  |
| 3 | Meditat* or Behavioral therap* or Psychotherap* or Psychodynamic therap* or Humanistic therap* or Integrative therap* or Systemic therap* or Exercise or Physical activity or Phototherapy or Light therap* or Yoga or Tai Ji or Tai-Ji or Tai Chi or Tai-Chi or Qigong or Qi Gong or Qi-Gong or S-adenosylmethionine or Omega-3 or Omega 3 or Herbal remed* or Herbal therap* or Hypericum Acupuncture |  |
| 4 | 1 and 2 and 3 | 2268 |
| **Canadian Ophthalmological Society** |  |  |
| 1 | depression | 0 |
| **American Academy of Ophthalmology** |  |  |
| 1 | depression | 1 |
| **Association for Vision in Research and Ophthalmology** |  |  |
| 1 | depression | 0 |

## Appendix 2. Brief descriptions of the interventions/exposures in the included studies

PST: PST is a cognitive-behavioral intervention that equips patients with the attitudes and skills to form solutions to everyday problems and in doing so, improve functioning.

SM: SM provide patients with practical skills and knowledge to play an active role in the management of their medical condition and its associated symptoms. Practical skills involve improving their independence. The cognitive component of SM involves teaching patients about their illness.

LVR: LVR is a multidisciplinary intervention led by an ophthalmologist to provide visually impaired individuals with the skills and tools to optimize their remaining visual capacity. This is accomplished through devices that either magnify their remaining vision or alter their environment to accommodate their visual impairment.

Optical and adaptive devices: Optical devices including magnifiers, telescopes, and specialized sun wear are prescribed by an ophthalmologist or optometrist and used to aid in near- and distance-reading tasks. Adaptive devices are likewise prescribed by a low vision professional and include such tools as large print text, talking appliances, and signature guide. These tools are used to assist in the completion of day-to-day tasks.

BA: BA is an intervention developed to treat depression by encouraging patients to engage in valued and rewarding activities that are self-reinforcing.

REBT: REBT is based on the theory that negative emotions arise from the irrational beliefs of the individual as opposed to any given event. As such, REBT teaches participants to practice logical thinking that is more flexible and less extreme.

Emotion-focused or problem-focused therapy: Emotion-focused therapy encourages participants to express and deal with negative emotions regarding AMD while problem-focused therapy encourages participants to discuss methods of addressing practical problems in their lives.

CBT: CBT is a form of talk therapy rooted in the theory that people’s emotions and behaviors are the result of their perceptions of an event as opposed to any given event in and of itself.

Mindfulness based music therapy: Mindfulness based music therapy combines components of both mindfulness therapy and music therapy. Mindfulness therapy involves directing one’s attention to their own sensation through various breathing and movement exercises. Music therapy involves exercises that include listening to various sounds and songs. Taken together, mindfulness-based music therapy involves performing mindfulness exercises while listening to various sounds and songs with the objective of deepening one’s awareness and acceptance of their internal state.

Alternate nostril breathing exercises: Alternate nostril breathing exercises involve breathing with the alternate occlusion of the left and right nostrils.

Written and audio tools: The written and audio tools that Kamga et al. (2017) administered was comprised of 3 main components.(55) Written and audio versions of the Antidepressant Skills Workbook taught practical skills to participants.(55) A Mood Monitoring Tool was administered as well.(55) These 2 components of the intervention were grounded in principles of CBT.(55) The third component of the intervention consisted of a DVD on depression.(55)

Motivational psychological nursing: Motivational psychological nursing is an intervention administered by nurses with the objective of creating a positive mood among patients to equip them with an optimistic outlook when managing their medical condition.

Expressive writing: Expressive writing is an intervention that enables participants to process emotions and experiences from past traumatic events by writing about them.

Alexander technique: The Alexander technique aims to bring awareness to maladaptive movement patterns and improve physical coordination by using verbal feedback and manual instruction.

Stepped care: In a stepped care model of treatment, patients are advanced to progressively more intensive stages of care based on their need.

Acupuncture: Acupuncture involves the insertion of metal needles into specific sites of the body to relieve pain.

